# Determinants of health as predictors for differential antibody responses following SARS-CoV-2 primary and booster vaccination in an at-risk, longitudinal cohort

**DOI:** 10.1101/2023.09.25.23296114

**Authors:** Felipe Echeverri Tribin, Erin Williams, Valeska Testamarck, Juan Manuel Carreño, Dominika Bielak, Temima Yellin, Florian Krammer, Michael Hoffer, Suresh Pallikkuth, Savita Pahwa

**Affiliations:** Department of Otolaryngology, University of Miami Miller School of Medicine, Miami, Florida, 33136 USA; Department of Microbiology and Immunology, University of Miami Miller School of Medicine, Miami, Florida, 33146, USA; Department of Biomedical Engineering, University of Miami, Miami, Florida, 33136, USA; University of Miami Miller School of Medicine, Miami, Florida, 33136, USA; Indiana University School of Medicine, Indianapolis, IN 46202, USA; Department of Neurological Surgery, University of Miami, Miller School of Medicine, Miami, Florida, 33136, USA; Department of Microbiology, Icahn School of Medicine at Mount Sinai, New York, New York, 10029, USA; Center for Vaccine Research and Pandemic Preparedness (C-VaRPP), Icahn School of Medicine at Mount Sinai, New York, NY, USA; Department of Pathology, Molecular and Cell-Based Medicine, Icahn School of Medicine at Mount Sinai, New York, NY, USA

**Keywords:** “SARS-CoV-2”, “primary vaccination”, “booster vaccination”, “humoral immunogenicity”, “breakthrough”, “cross-sectional analysis”, “linear mixed-effects models”

## Abstract

Post vaccine immunity following COVID-19 mRNA vaccination may be driven by extrinsic, or controllable and intrinsic, or inherent health factors. Thus, we investigated the effects of extrinsic and intrinsic on the peak antibody response following COVID-19 primary vaccination and on the trajectory of peak antibody magnitude and durability over time. Participants in a longitudinal cohort attended visits every 3 months for up to 2 years following enrollment. At baseline, participants provided information on their demographics, recreational behaviors, and comorbid health conditions which guided our model selection process. Blood samples were collected for serum processing and spike antibody testing at each visit. Cross-sectional and longitudinal models (linear-mixed effects models) were generated to assess the relationship between selected intrinsic and extrinsic health factors on peak antibody following vaccination and to determine the influence of these predictors on antibody over time. Following cross-sectional analysis, we observed higher peak antibody titers after primary vaccination in females, those who reported recreational drug use, younger age, and prior COVID-19 history. Following booster vaccination, females and Hispanics had higher peak titers after the 3^rd^ and 4^th^ doses, respectively. Longitudinal models demonstrated that Moderna mRNA-1273 recipients, females, and those previously vaccinated had increased peak titers over time. Moreover, drug users and half-dose Moderna mRNA-1273 recipients had higher peak antibody titers over time following the first booster, while no predictive factors significantly affected post-second booster antibody responses. Overall, both intrinsic and extrinsic health factors play a significant role in shaping humoral immunogenicity after initial vaccination and the first booster. The absence of predictive factors for second booster immunogenicity suggests a more robust and consistent immune response after the second booster vaccine administration.

## Introduction

Severe acute respiratory syndrome coronavirus-2 (SARS-CoV-2) infection has had rapid and widespread international effects since its onset in late December 2019 (1–5). Aside from the socioeconomic burdens and turmoil generated from its spread, early access to health interventions and prophylactic measures for the virus were limited and ineffective at controlling the spread. Instead, government-imposed lockdowns and social distancing were utilized as preventative measures (4), but due to high transmissibility and virulence the U.S. Food and Drug Administration (FDA) granted emergency use authorization (EUA) for nearly 400 coronavirus disease-19 (COVID-19) related medical products in the United States, including diagnostic tests, medical devices, drugs, and vaccines (4, 6–8), and helped distribute vaccines worldwide.

Prior work has shown that post-vaccine immunity is driven by various factors, including prior SARS-CoV-2 infection, vaccine manufacturer, heterologous booster administration, time since vaccination, and the magnitude of peak antibody titers after vaccination (9–18). Furthermore, both systemic and local symptoms following SARS-CoV-2 infection and vaccination have been associated with higher peak antibody responses following primary vaccination (5). Overall, COVID-19 vaccines and boosters have been demonstrated to safely curb infection and severe SARS-CoV-2 outcomes, but more work is needed to understand drivers for post-vaccination heterogeneity. The effect of intrinsic health factors (IHFs) and extrinsic health factors (EHFs) on COVID-19 vaccine immunogenicity is unknown. IHFs are characteristics that relate to an individual and cannot be changed or manipulated (e.g., age, biological sex, or race), while EHFs relate to health comorbidities and behaviors (e.g., diet, smoking, or drug consumption) that can be altered or controlled.

Accounting for IHFs and EHFs when evaluating post-vaccine immunogenicity in large populations could result in more personalized analyses from a public health approach (5). For example, prior research has shown that individuals with diabetes mellitus and obesity exhibit dampened immune responses to COVID-19 and hepatitis B vaccines, compared to controls (19, 20). Apart from chronic disease, age, and biological sex, human leukocyte antigen types have also been identified as factors that can cause heterogeneous immune reactions to vaccination due to senescence and variable cytokine production among different groups (21–24). Despite these findings, the evaluation of IHFs and EHFs have yet to be determined in a longitudinal cohort following COVID-19 vaccination. Thus, the purpose of this study is to investigate the effects of IHFs and EHFs on peak antibody (Ab) titers following COVID-19 primary vaccination (PV) and booster vaccination (BV), and to investigate how these factors influence Ab magnitude and trajectory over time.

## Methods

### Study design

Participants were enrolled in our Institutional Review Board (IRB) approved (#20201026), longitudinal, observational SARS-CoV-2 immunity study (n=228) known as “CITY” (**C**OVID-19 **I**mmuni**TY** study) as part of the PARIS/SPARTA study (25). Recruitment for the study began in October 2020 and ended in February 2023. The study was conducted at the University of Miami Miller School of Medicine, and study subjects participated in visits every 3 months for a total of 2 years from the time of enrollment. The primary objective of the CITY study was to characterize differential antibody kinetics among SARS-CoV-2 uninfected and infected individuals in a high-risk, ethnically diverse cohort. The “high-risk” designation for inclusion referred to hazardous occupational exposure (e.g., healthcare workers) but also to advanced age or other sociodemographic characteristics known to increase risk of SARS-CoV-2 morbidity and mortality. Following written informed consent, participants provided demographic details to include social and lifestyle habits and relevant past medical history that would preclude them to more severe SARS-CoV-2 infection-related outcomes. They also provided details regarding their past medical history to include conditions like hypertension, hypercholesterolemia, previous SARS-CoV-2 infection, diabetes, and COVID-19 vaccine manufacturer, lifestyle factors like alcohol or drug consumption, and other details such as age, biological sex, educational level, employment status, marital status, race, ethnicity, and sexual orientation. Those who suffered from SARS-CoV-2 infection with a documented positive nucleic acid amplification test (NAAT+) prior to entry provided evidence details regarding their past SARS-CoV-2 infection associated symptoms during the baseline visit to the study team.

At all regularly scheduled visits, participants prospectively answered symptom questionnaires to screen for new or recent SARS-CoV-2 infection, and blood samples were collected for serum and peripheral blood mononuclear cell (PBMC) processing. Plasma was stored at -80°C and PBMCs were cryopreserved in liquid N_2_ (26). All participants agreed to sample banking for future research use (Fig 1).

**Fig 1.**
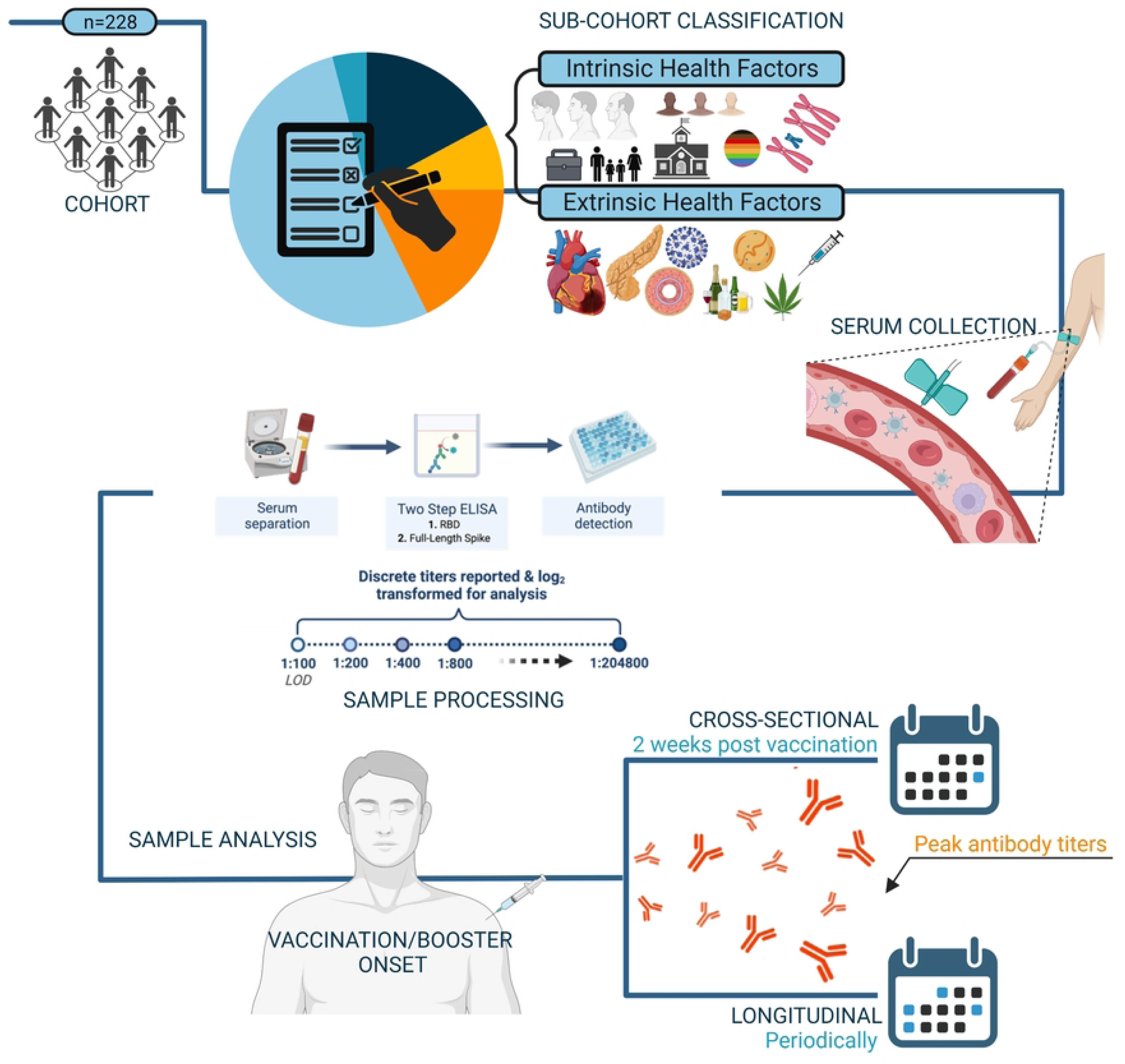
Overview of Experimental Methods. Classification of intrinsic and extrinsic health factors were determined by participant responses gathered during baseline questionnaires. Blood samples were collected at regular intervals, followed by processing and ELISA-mediated antibody detection with discrete titers reported from 1:100 to 1:204800. Sample analysis was achieved through cross-sectional and longitudinal analyses. Created with BioRender.com

### SARS-CoV-2 ELISAs

SARS-CoV-2 enzyme-linked immunosorbent assays (ELISAs) were performed according to a well described assay developed by the Icahn School of Medicine at Mount Sinai (27, 28). 96-well plates were collected at 4°C with wild-type Wuhan SARS-CoV-2 spike protein (2 mg/ml) solution and incubated overnight. Plates were blocked with 3% non-fat milk prepared in phosphate buffered saline with 0.1% Tween 20 (PBST) and incubated at room temperature for an hour. After blocking, serial dilutions of heat-inactivated serum samples were added to the plates and incubated for two hours at room temperature. Plates were washed in triplicate with 0.1 % PBST and 50 µl of a 1:3,000 dilution of goat anti-human immunoglobulin G-horseradish peroxide (HRP) conjugated secondary antibody was added. After an hour of incubation, 100 µl *o-*phenylenediamine dihydrochloride (SIGMAFASTTM-OPD, *Sigma Aldrich*) solution was added to each well for 10 minutes. The reaction was stopped by the addition of 50 µl of 3 M hydrochloric acid at each well. The optical density at 490 nm (OD490) was measured using a Synergy 4 (BioTek) plate reader. The background value (OD490 = 0.15) allowed to report discrete antibody endpoint titers in the values of 1:100, 1:200, 1:400, 1:800, 1:1600, 1:3200, 1:6400, 1:12800, 1:25600, 1:51200, 1:102400, and 1:204800. The limit of detection was set at 1:100.

### Statistical analyses

For all analyses, regardless of the statistical approach, Ab titers were log_2_ transformed. Intrinsic and extrinsic factors (Table 1) were included in the cross-sectional and longitudinal analyses detailed below.

**Table 1.**
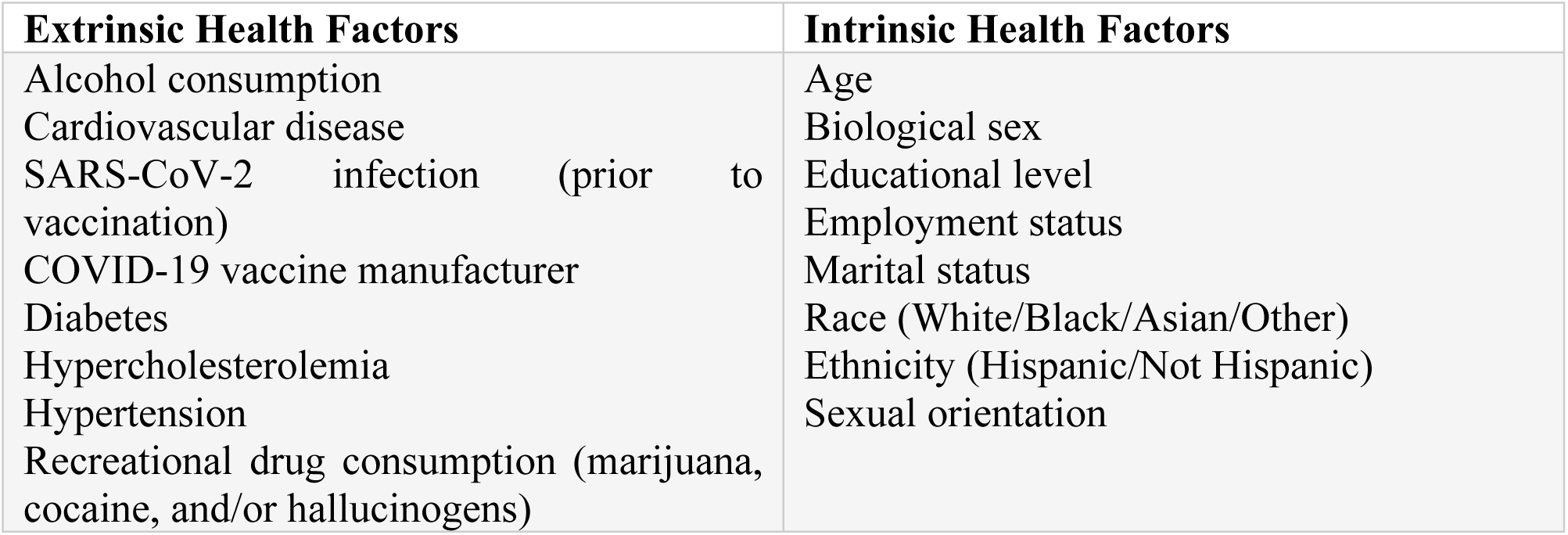
Observed Extrinsic and Intrinsic Health Factors Selected as Covariates.

#### Cross-sectional analysis

For the cross-sectional analysis, all respondents had participated in additional visits with associated Ab titers measured within 2 weeks [± 5 days] following primary and booster vaccination. Cross-sectional analysis was conducted separately for post-vaccine timepoints following PV, BV1, and BV2. We included only those (n = 118) who had received their primary vaccination (i.e., two doses of Pfizer BNT162b2 or two doses of Moderna mRNA-1273) after their inclusion into the study and strictly within the two-dose schedule recommended by the Centers for Disease Control and Prevention (CDC). This approach minimized potential discrepancies in vaccination effects based on administration time.

The underlying assumptions for parametric tests (e.g., tests for homogeneity of variance and normality) were conducted for Ab titers among IHF and EHF groups following each post-vaccination sub cohort of interest (i.e., PV1, BV1, and BV2) prior to conducting hypothesis testing. Differences in mean Ab were analyzed using parametric (one-way analysis of variance (ANOVA), Student’s t-test) or non-parametric (Games-Howell, Welch’s t-test) tests, based on Levene’s test results. Lastly, Pearson’s correlation was used to examine the correlation between age and peak Ab titer. All statistical analyses were conducted via Minitab (*Minitab® 21.3.1*) at a significance level of α = 0.05, and tests for groups with sample sizes <2 were excluded from analysis.

### Longitudinal analysis

The selection of covariates of interest for the longitudinal analysis was informed by the aforementioned cross-sectional analysis conducted at post-primary vaccination timepoints. Preliminary analyses utilized a range of potential predictors to evaluate their association with the response variable, log_2_ Ab titers over time. From this subset, only those predictors that exhibited a significant association log_2_ Ab were carried forward as covariates of interest for the longitudinal analysis. This method ensured that the longitudinal model was informed by empirically significant predictors, thus increasing the potential explanatory power and relevance of the model to the observed data. Other proven covariates of interest (5), including COVID-19 vaccine manufacturer, were included during the model generation phase as well.

Following the above, linear mixed-effects models were constructed to investigate the growth curve of log_2_ antibody titers over time following primary vaccination (Dose 1 and 2), as well as booster vaccination (Dose 3 and Dose 4) utilizing the *nlme* and *lme4* package in R Studio. The models were designed to incorporate both fixed and random effects, with the fixed effects representing the average population response and the random effects capturing subject-specific deviations. The process of model generation was initiated by constructing a maximal model, where the time since vaccination – represented as a continuous variable – was incorporated as additional polynomial terms to account for potential non-linear growth patterns. This maximal model considered the most complex growth trajectory from the outset.

To account for the repeated measures structure of the data, various correlation structures including compound symmetry, first-order autoregressive, and general autoregressive were compared. Model comparison was performed using the Akaike Information Criterion (AIC) and the Bayesian Information Criterion (BIC), with the model yielding the lowest values being selected as the optimal model.

## Results

### Cohort characteristics

#### Extrinsic health factors - summary

As seen in Table 1, among those enrolled (n = 228), the average age in the “CITY” cohort was 48.6 ± 16.56 years, with 66 (34.74%) participants classified as young, 84 (44.21%) as middle aged, and 40 (21.05%) as elderly (n = 190).

Table 2 displays a summary table for the cohort extrinsic health factors. At baseline, n = 180 (78.95%) participants self-reported as alcohol consumers and 47 (20.61%) did not (n = 227). There were 9 (3.96%) participants with a history of cardiovascular disease (CVD), 216 (95.16%) with no history of CVD, and 2 (0.88%) that were unsure (n = 227). Also, 9 (3.96%) participants were diabetic, while 215 (94.72%) were not, and 3 (1.32%) were unsure (n = 227). Moreover, 16 (7.08%) participants were recreational drug users and 210 (92.92%) were not (n = 226). Among the cohort, 39 (17.18%) of the participants had hypercholesterolemia, while 173 (76.21%) did not, and 14 (6.17%) were unsure (n = 227). Lastly, 37 (16.30%) participants reported hypertension, 185 (81.50%) did not, and 5 (2.20%) were unsure (n = 227).

**Table 2.**
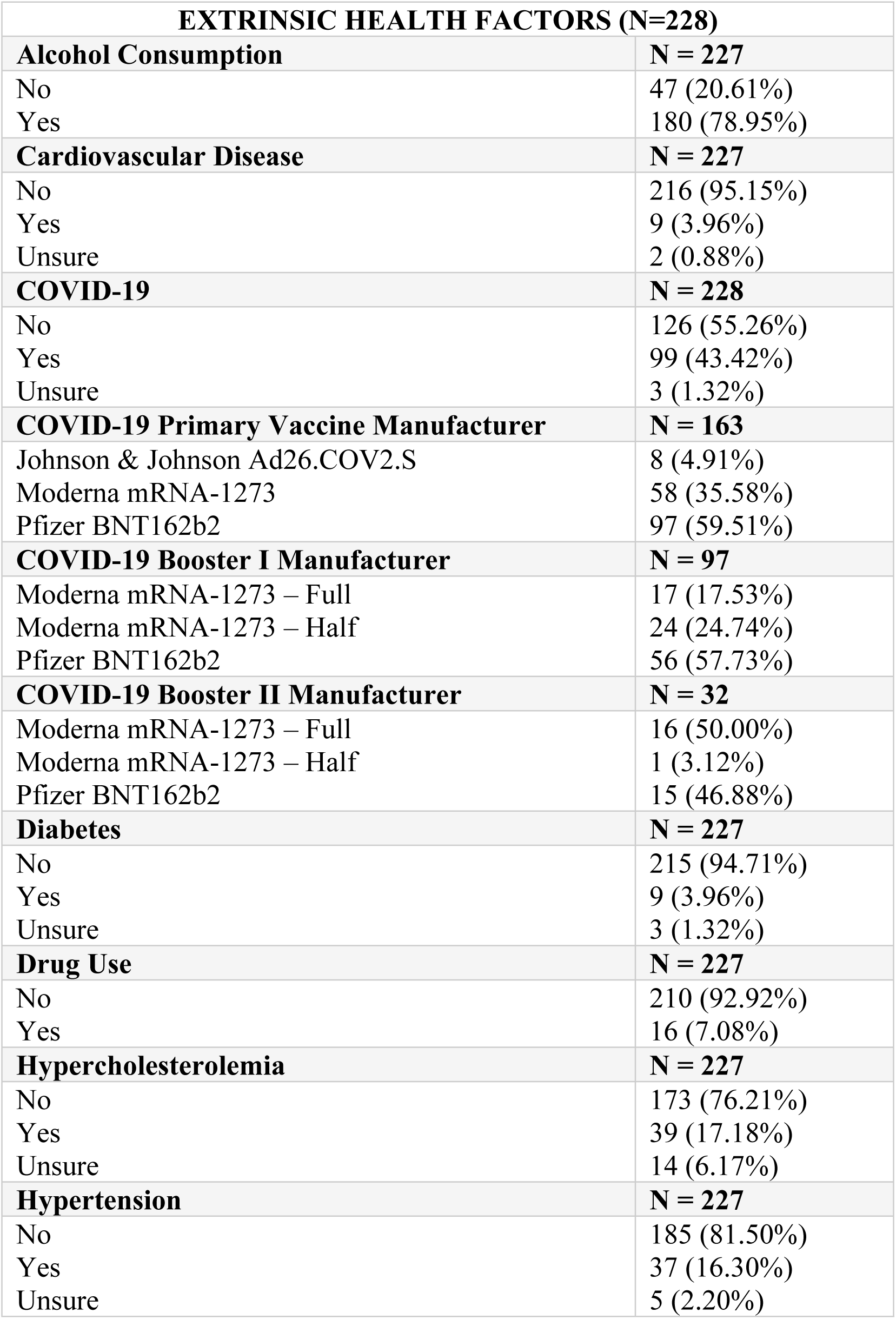
Cohort Extrinsic Health Factor Summary Table.

Ninety-nine (43.42%) participants indicated previous SARS-CoV-2 infection at entry, while 126 (55.26%) did not, and 3 (1.32%) were unsure (n=228). Of the 163 participants in the cohort that received PV, 8 (4.91%) received Johnson & Johnson Ad26.COV2.S, 58 (35.58%) received Moderna mRNA-1273, and 97 (59.51%) received Pfizer BNT162b2. Of the 97 participants that received the first booster (BV1), 17 (17.53%) received the full Moderna mRNA-1273 dose, 24 (24.74%) received the half-dose Moderna mRNA-1273, and 56 (57.73%) received Pfizer BNT162b2. Of those 32 participants that received a second booster (BV2), 16 (50%) received a full Moderna mRNA-1273 dose, 1 (3.12%) received the Moderna mRNA-1273 half dose, and 15 (46.88%) received a Pfizer BNT162b2 booster. No bivalent boosters were included in the analysis.

#### Intrinsic health factors - summary

Table 3 displays a summary table for the cohort intrinsic health factors. One hundred and thirty-seven (60.00%) participants were female and 91 (40.00%) were male (n = 228). Furthermore, it was reported that 23 (10.10%) participants had an associate degree or a technical degree, 49 (21.49%) had a bachelor’s degree, 12 (5.26%) had a high school diploma or equivalent, 3 (1.32%) had less than a high school diploma, 111 (48.68%) had a master’s degree or higher, and 29 (12.72%) had other educational levels (n = 227).

**Table 3.** Cohort Intrinsic Health Factor Summary Table.

| **INTRINSIC HEALTH FACTORS (N = 228)** |  |
| --- | --- |
| **Age** | **N = 190** |
| <40 – Young | 66 (34.74%) |
| 40-65 – Middle | 84 (44.21%) |
| >65 – Elderly | 40 (21.05%) |
| **Biological Sex** | **N = 228** |
| Female | 137 (60.09%) |
| Male | 91 (39.91%) |
| **Education Level** | **N = 227** |
| Associate degree or technical degree | 23 (10.13%) |
| Bachelor's degree | 49 (21.59%) |
| High school diploma or equivalent | 12 (5.29%) |
| Master's degree or higher | 111 (48.90%) |
| Other | 29 (12.78%) |
| **Employment Status** | **N = 226** |
| Disabled | 2 (0.88%) |
| Employed | 188 (83.19%) |
| Retired | 24 (10.62%) |
| Unemployed | 12 (5.31%) |
| **Hispanic/Non-Hispanic** | **N = 228** |
| Hispanic | 90 (39.47%) |
| Non-Hispanic | 138 (60.53%) |
| **Marital Status** | **N = 228** |
| Divorced | 19 (8.33%) |
| Domestic partnership | 11 (4.82%) |
| Married | 113 (49.56%) |
| Single | 79 (34.65%) |
| Widowed | 6 (2.64%) |
| **Race** | **N = 228** |
| Asian | 12 (5.26%) |
| Black | 12 (5.26%) |
| Other | 21 (9.21%) |
| White | 183 (80.27%) |
| **Sexual Orientation** | **N = 228** |
| Bisexual | 3 (1.32%) |
| Gay | 15 (6.58%) |
| Heterosexual | 206 (90.34%) |
| Lesbian | 3 (1.32%) |
| Other | 1 (0.44%) |

Additionally, 2 (0.87%) participants were disabled, 188 (82.46%) were employed, 24 (10.54%) were retired, and 12 (5.26%) were unemployed (n=226). Nineteen (8.33%) were divorced, 11 (4.82%) participated in a domestic partnership, 113 (49.57%) were married, 79 (34.65%) were single, and 6 (2.63%) were widowed (n = 228). In addition, 12 (5.26%) participants identified as Asian, 12 (5.26%) as African American, 183 (80.27%) as White, and 21 (9.21%) as part of another race (n = 228). Ninety (39.47%) participants identified as Hispanic, and 138 (60.53%) participants identified as non-Hispanic. Lastly, 3 (1.32%) participants identified as bisexual, 15 (6.58%) as gay, 206 (90.35%) as heterosexual, 3 (1.32%) as lesbian, and 1 (0.43%) as another sexual orientation (n = 228).

### Primary vaccination

Overall, 148 participants were included in the PV sub-cohort, with an average peak titer of 13.29 ± 1.67 following vaccination. Table 4 details all significant differences observed across extrinsic and intrinsic health factor covariates in the PV sub-cohort or that had significance in the BV1 and BV2 sub-cohorts. Other measures examined for the purpose of this analysis are documented in S1 Table.

**Table 4.**
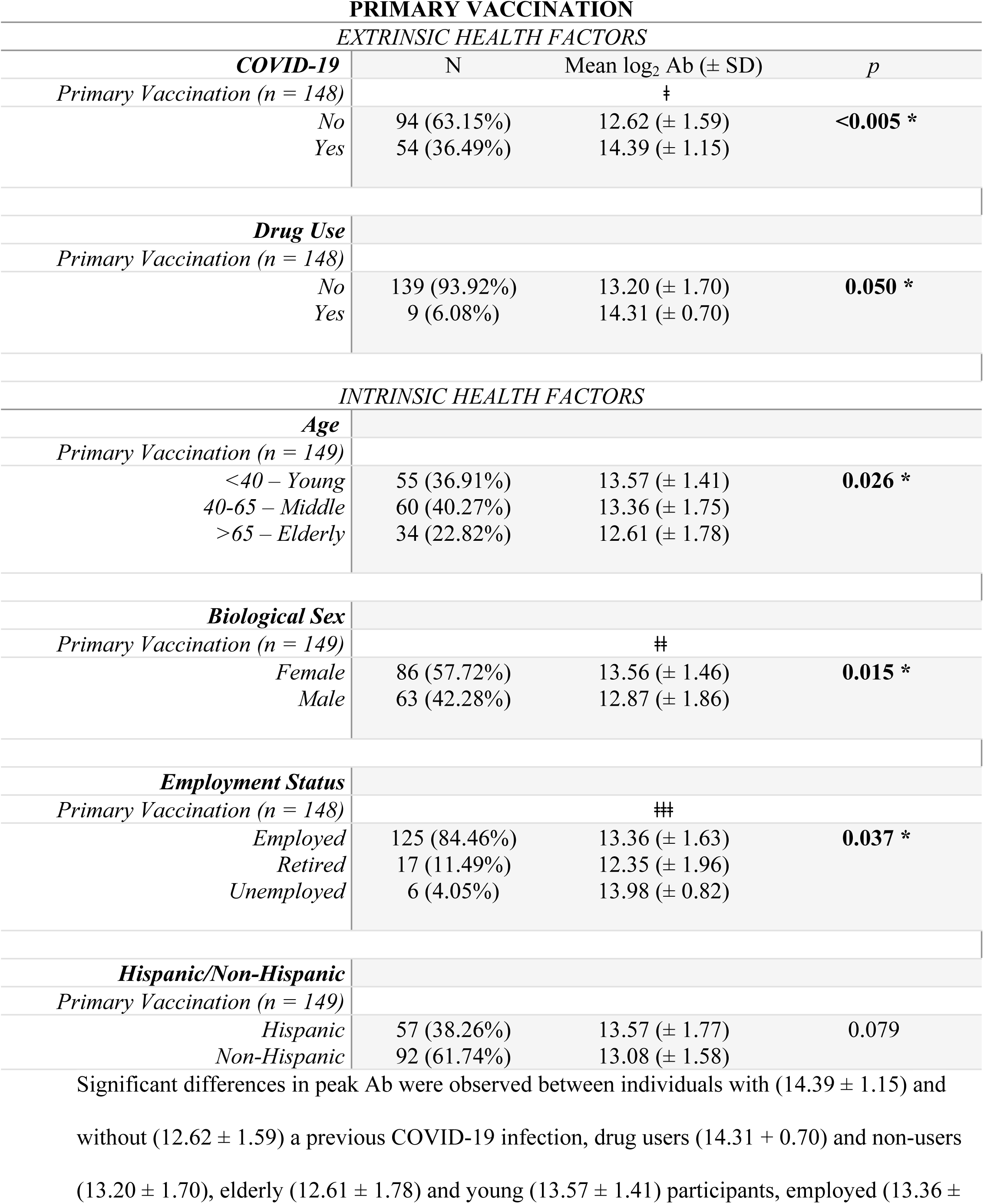
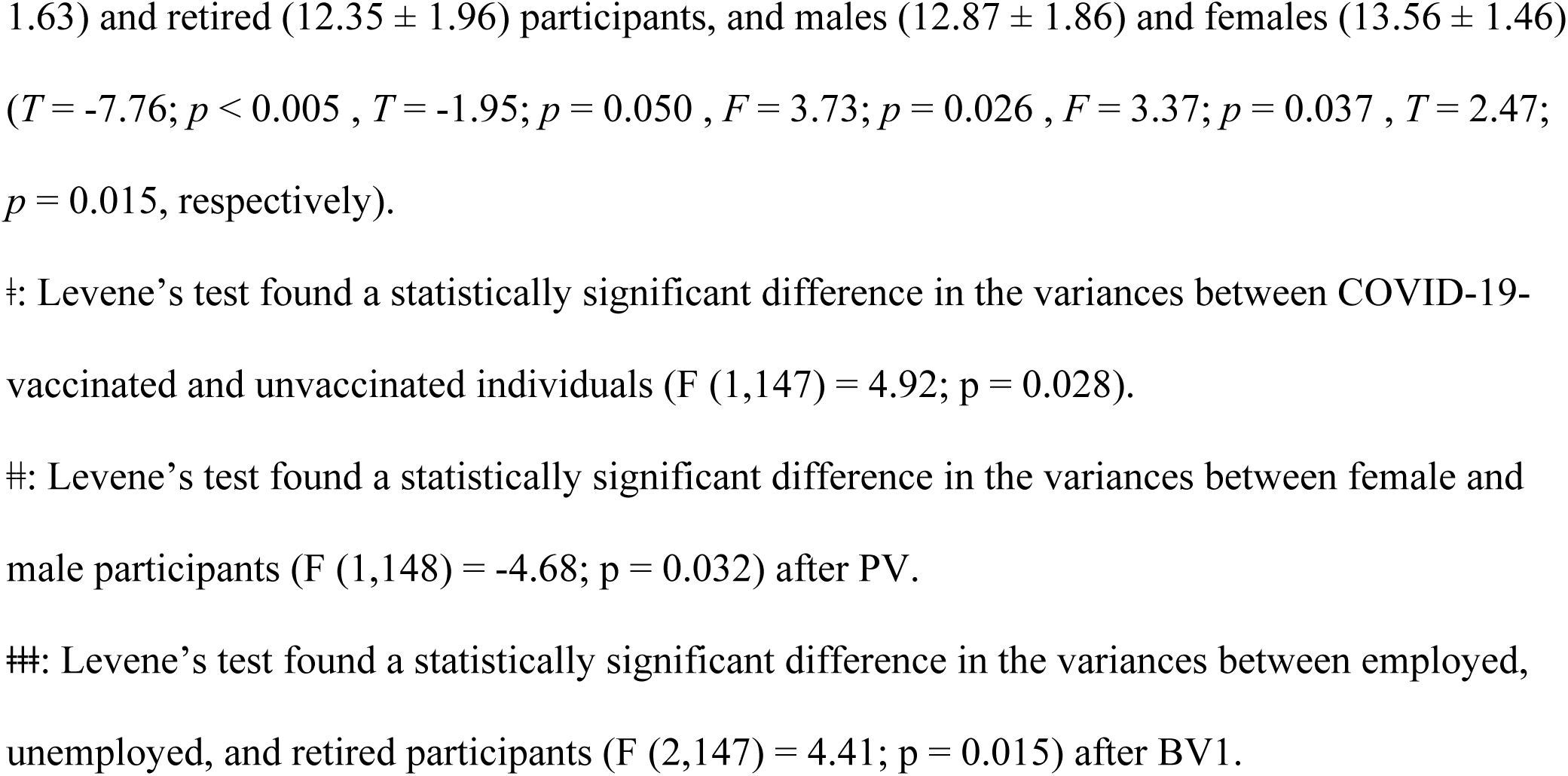
Mean Log_2_ Ab Titers in Primary Vaccination Sub-Cohort with Varying Extrinsic and Intrinsic Health Factors.

#### Extrinsic health factors

Five (3.42%) individuals included in the PV sub-cohort had CVD. while the remaining 141 (96.58%) did not. The mean log_2_ Ab of participants with CVD (12.84 ± 3.11) and without CVD (13.29 ± 1.59) were not statistically different (*T* = 0.59; *p* = 0.555). Four who received PV (2.72%) had diabetes and 143 (97.28%) did not. The mean log_2_ Ab of participants with diabetes (12.89 ± 2.87) and participants without diabetes (13.31 ± 1.61) were not statistically different (*T* = 0.50; *p* = 0.619). Twenty-seven (19.42%) had hypercholesterolemia while 112 (80.58%) did not. The mean log_2_ Ab of participants with hypercholesterolemia (13.01 ± 1.96) and without hypercholesterolemia (13.31 ± 1.60) were not statistically different (*T* = 0.83; *p* = 0.405). Furthermore, 22 (15.07%) had hypertension and 124 (84.93%) did not have hypertension. The variances of log_2_ Ab between participants with hypertension and participants without hypertension were statistically different, (*F* (1,145) = 8.15; *p* = 0.005) and indicated that individuals without hypertension had lower and more homogenous Ab response after PV. However, the mean log_2_ Ab of participants with hypertension (12.69 ± 2.36) and participants without hypertension (13.34 ± 1.50) were not statistically different (*T* = 1.24; *p* = 0.225).

Fifty-four (36.49%) of those that received PV reported prior SARS-CoV-2 infection at study entry and 94 (63.51%) did not. The variances of log_2_ Ab between participants with a history of COVID-19 and no history of COVID-19 were statistically different (*F* (*1, 147*) = 4.92; *p* = 0.028), as were the peak responses among participants with a COVID-19 history (14.39 ± 1.15) and no history of COVID-19 (12.62 ± 1.59) (*T* = - 7.76; *p* < 0.005). Furthermore, 149 participants received PV, where 54 (36.24%) received a Moderna vaccine and 95 received a Pfizer BNT162b2. The mean log_2_ Ab of participants with the Moderna mRNA-1273 vaccine (13.50 ± 1.71) and the Pfizer BNT162b2 vaccine (13.14 ± 1.64) were not statistically different, as shown in Fig 2 (*T* = 1.26; *p* = 0.211). Of the 148 participants who received PV, 119 (80.41%) were alcohol consumers and 29 (19.59%) were not alcohol consumers. The mean log_2_ Ab of alcohol consumers (13.39 ± 1.58) and non-alcohol consumers (12.75 ±1.97) were not significantly different (*T* = - 1.87; *p* = 0.063) after PV. Moreover, of the 148 participants that received PV, 9 (6.08%) were recreational drug users and 139 (93.92%) were not. The mean log_2_ Ab of recreational drug users (14.31 ± 0.70) and non-users (13.20 ± 1.70) were statistically different (*T* = -1.95; *p* = 0.050) after PV.

**Fig 2.**
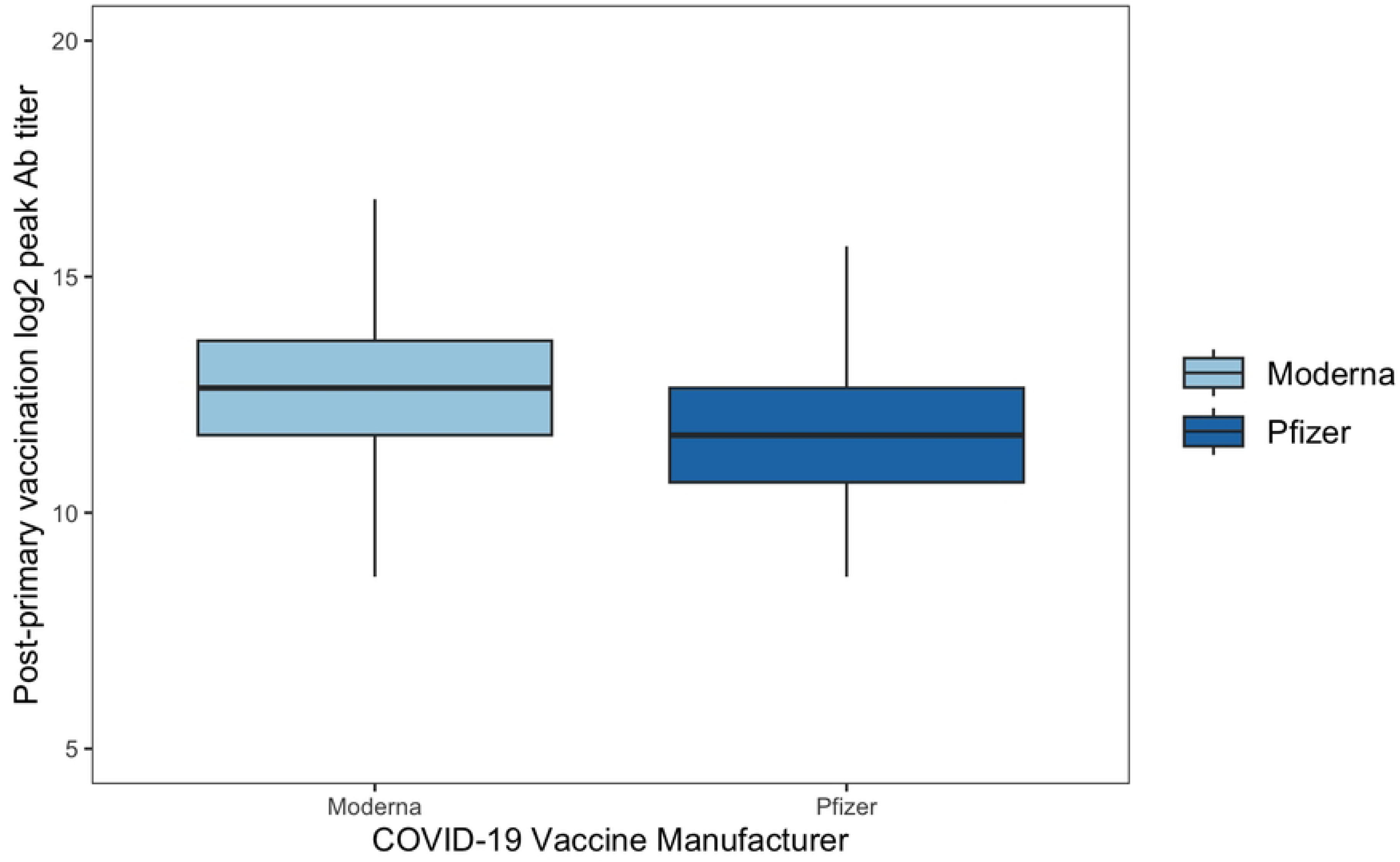
Post-Primary Vaccination Mean Log_2_ Ab Across COVID Vaccine Groups. There was no significant difference (T=1.26, p=0.211) between mean log_2_ Ab between participants who received Moderna mRNA-1273 (13.50 ± 1.71) or Pfizer BNT162b2 vaccines (13.14 ± 1.64) for their primary vaccination after 2 weeks of vaccination onset.

#### Intrinsic health factors

Of the 149 participants that received PV, 55 (36.91%) of them were young, 60 (40.27%) were middle aged, and 34 (22.82%) were elderly. The mean log_2_ Ab of low age participants (13.57 ± 1.41; Tukey = A) was statistically different (*Fig 3*) than elderly participants (12.61 ± 1.78 ; *Tukey* = B), but not middle-aged participants (13.36 ± 1.75 ; *Tukey* = A, B) after PV (*F* = 3.73 ; *p* = 0.026). Pearson correlation demonstrated a negative correlation between age and log_2_ Ab after PV (*r* = -0.206; *p* = 0.012). Additionally, 86 (57.72%) participants were female and 63 (42.28%) were male. The variances of the log_2_ Ab between female and male participants were significantly different (*F* (1,148) = -4.68; *p* = 0.032) after PV. Furthermore, male participants exhibited a lower (12.87 ± 12.87) was statistically lower (*T* = 2.47; *p* = 0.015) than that of female (13.56 ± 1.46) participants after PV (*Fig 4*).

**Fig 3.**
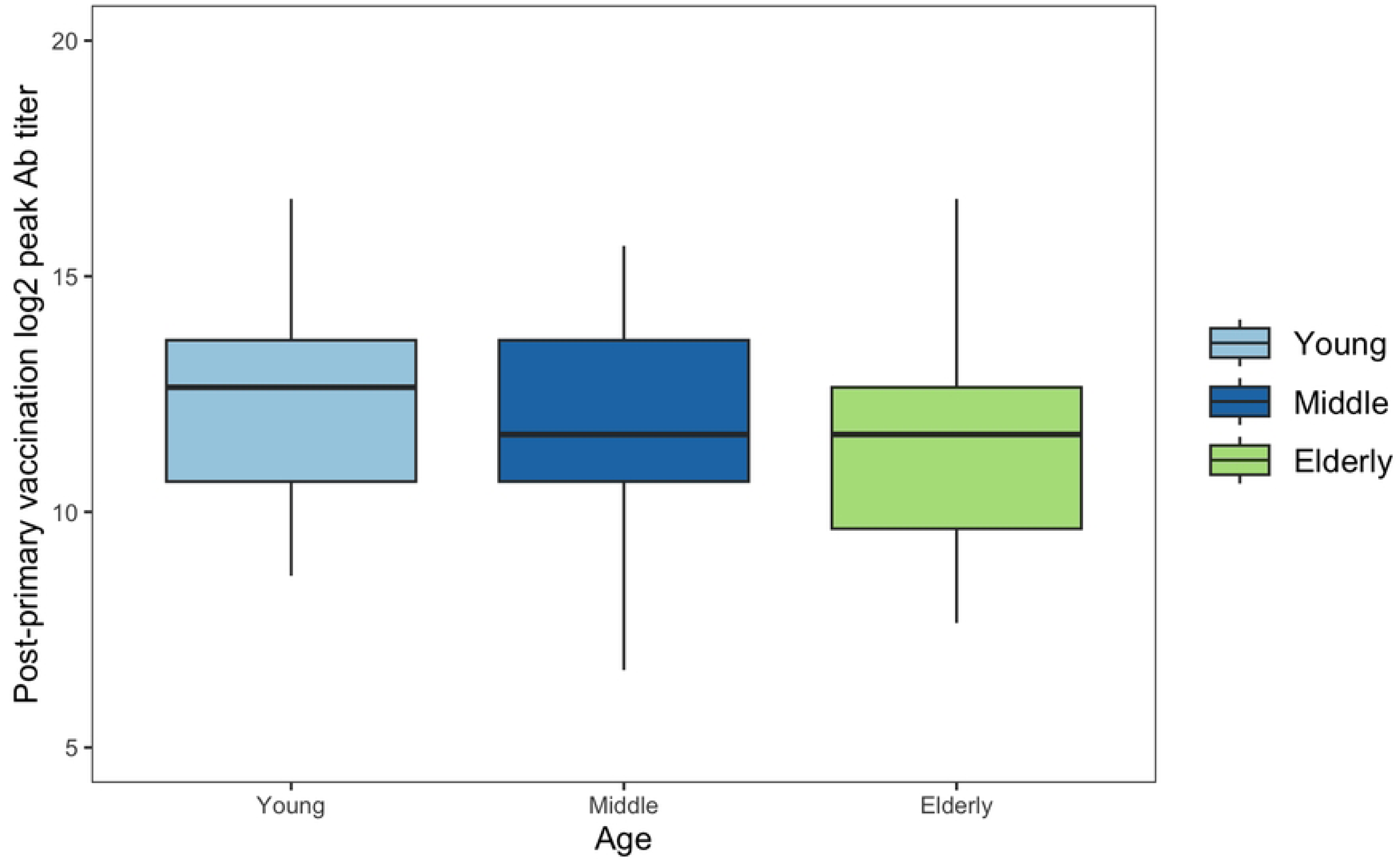
Post-Primary Vaccination Means Log_2_ Ab Across Age Groups. The mean log2 Ab of low age participants (13.57 ± 1.41; Tukey = A) was statistically different than elderly participants (12.61 ± 1.78 ; Tukey = B), but not middle-aged participants (13.36 ± 1.75 ; Tukey = A, B) after primary vaccination (*F* = 3.73 ; *p* = 0.026).

**Fig 4.**
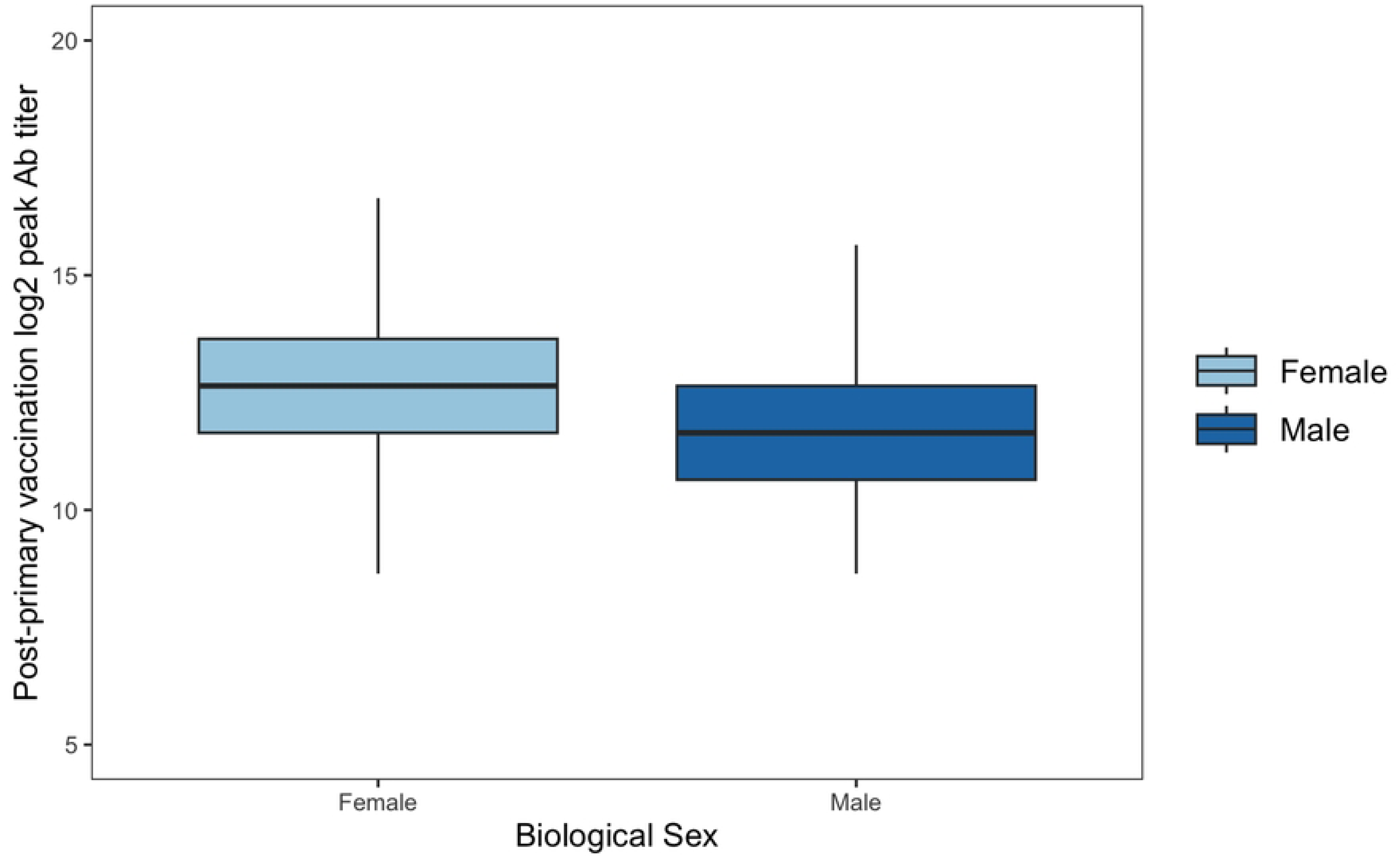
Post-Primary Vaccination Mean Log_2_ Ab Across Biological Sex Groups. The mean log_2_ Ab in male participants (12.87 ± 12.87) was statistically lower (T=2.47, p=0.015) than that of female (13.56 ± 1.46) participants after primary vaccination

Furthermore, of the 149 participants that received PV, 12 (8.05%) reached an education level of an associate degree or technical degree, 30 (20.13%) held a bachelor’s degree, 9 (6.04%) a high school diploma, 75 (50.34%) a master’s degree or higher, and 23 (15.44%) reported reaching an unlisted level. Peak titers were not associated with differences between any group (*F* = 1.87) ; (*p* = 0.120), including those with an associate degree or technical degree (13.56 ± 1.44 ; *Tukey* = A), a bachelor’s degree (13.68 ± 1.69 ; *Tukey* = A), a high school diploma (13.98 ± 1.58 ; *Tukey* = A), a master’s degree or higher (13.16 ± 1.64 ; *Tukey* = A), and other educational levels (12.64 ± 1.76 ; *Tukey* = A). One-hundred and twenty-five (84.46%) were employed, 17 (11.49%) were retired, and 6 (4.05%) were unemployed. The mean log_2_ Ab of employed (13.36 ± 1.63, *Tukey* = A), retired (12.35 ± 1.96; *Tukey* = A), and unemployed (13.98 ± 0.82, *Tukey* = A) participants were statistically different (*F* = 3.37 ; *p* = 0.037) after PV, though the authors speculate that this finding is due to a strong correlation between age and the retired group.

Twelve (8.05%) participants were divorced, 7 (4.70%) participated in a domestic partnership, 81 (54.36%) were married, 47 (31.54%) were single, and 2 (1.35%) were widowed. The post PV Ab between divorced (13.81 ± 1.24, *Tukey* = A), domestic partnership (13.07 ± 1.51, *Tukey* = A), married (13.04 ± 1.82, *Tukey* = A), and single (13.54 ± 1.49, *Tukey* = A) participants were not statistically different (*F* = 1.36; *p* = 0.257) after PV. Only 2 (1.34%) identified as bisexual, 10 (6.71%) identified as gay, 134 (89.94%) identified as heterosexual, 2 identified as lesbian (1.34%), and 1 (0.67%) identified as another sexual orientation. Furthermore, the mean log_2_ Ab between gay (13.34 ± 0.45) and heterosexual (13.24 ± 0.15) participants were not significantly different (*T* = 0.19; *p* = 0.853) after PV.

Also, of the 149 participants that received PV, 9 (6.04%) were Asian, 6 (4.03%) were African American, 14 (9.40%) identified under another race, and 120 (80.54%) were White, though the mean peak response (13.09 ± 1.88, *Tukey* = A; 13.98 ± 0.82, *Tukey* = A; 13.43 ± 1.37, *Tukey* = A; 13.23 ± 1.72, *Tukey* = A; respectively) were not statistically different (*F* = 0.46 ; *p* = 0.712) after PV. Moreover 57 (38.26%) participants identified as Hispanic, and 92 (61.74%) identified as non-Hispanic. The mean log_2_ Ab between Hispanic (13.57 ± 1.77) and non-Hispanic (13.08 ± 1.58) participants were not statistically different (*T* = 1.77; *p* = 0.079) after PV.

#### Longitudinal analysis following primary vaccination

A linear mixed-effects model (Fig 5) was employed to investigate the fixed effects of days since full vaccination (including linear, quadratic, and cubic time), COVID-19 vaccine manufacturer, COVID-19 status, and biological sex on the log-transformed titer levels (antibody), while accounting for random effects associated with each study participant. The fixed effects of linear, cubic, and quadratic time revealed that days since full vaccination (i.e., >14 days following the 2^nd^ dose) and its second and third powers were negatively associated with log_2_ antibody titers (*estimate* = -4.653e^-03; *F*(1,524) = 96.889; *p* = <0.001, *estimate* = -2.208e^-04; F(1,524) = 118.393; *p* <0.001, and *estimate* = 9.467e^-07; *F*(1,24) = 44.654; *p* < 0.001, respectively). Participants who had been vaccinated with Pfizer BNT162b2 had a lower antibody response over time compared to those who received the Moderna mRNA-1273 vaccine (*estimate* = -9884e^- 01; *F*(1,108) = 36.900; *p* < 0.001).

**Fig 5.**
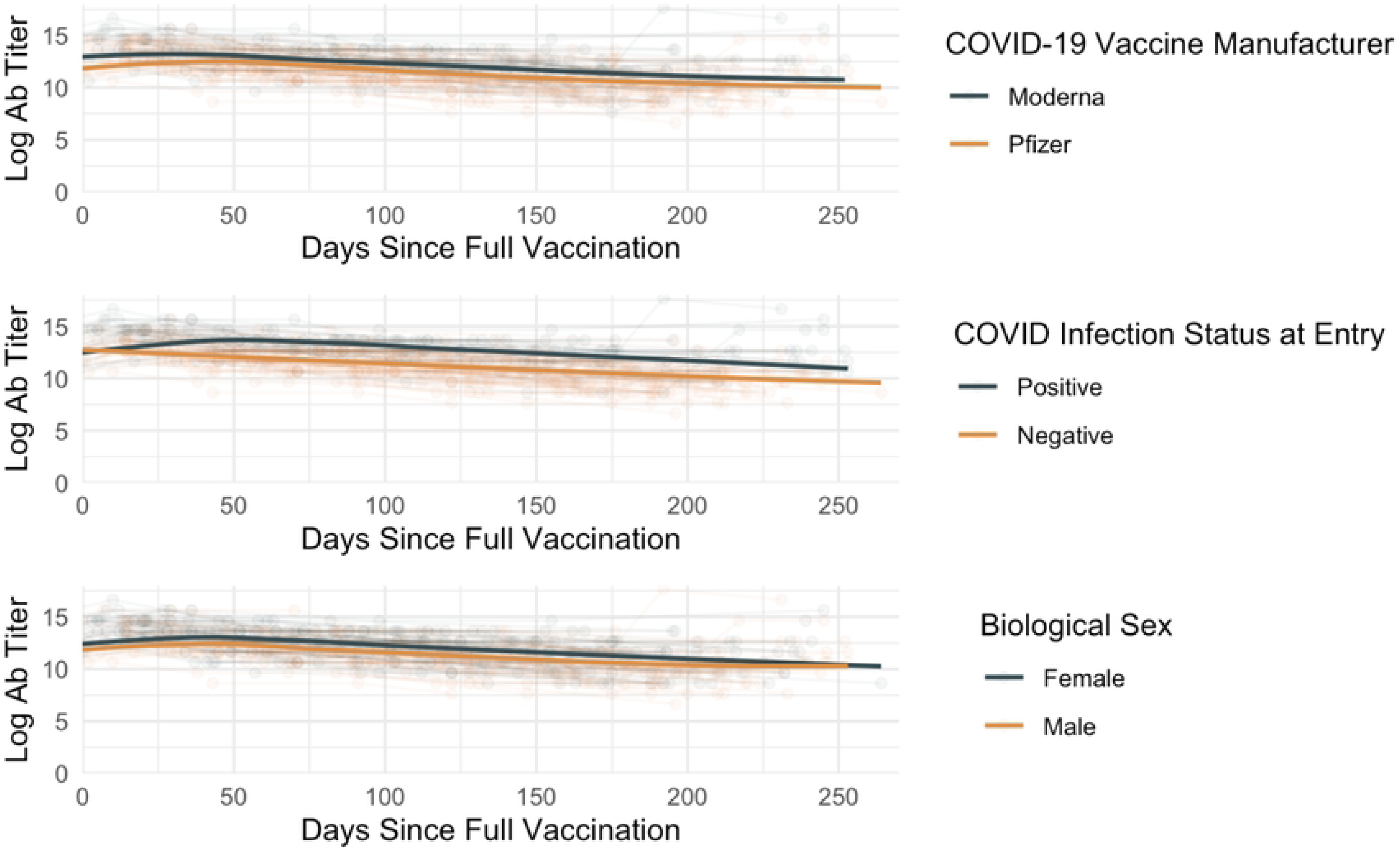
Linear Mixed-Effects Model for Investigation of Fixed Effects of Days Since Full Vaccination, Primary Covid-19 Vaccine Manufacturer, Covid-19 Status, And Biological Sex. Moderna mRNA-1273 recipients exhibited higher peak titers as well as more durable antibody responses when compared to Pfizer BNT162b2, as did study participants who were previously COVID-19 positive prior to vaccine recipient. Male participants, on average, demonstrated lower Ab responses when compared to female participants.

Additionally, COVID-19 positive participants had higher antibody levels compared to COVID-19 negative individuals (*estimate* = 1.464; *F*(1,108) = 64.917; *p* < 0.001), consistent with well-described trends for hybrid immunity (29). The analysis also suggested a difference in antibody levels based on biological sex, with males having lower levels compared to females (*estimate* = -3.759e^01; *F*(1,108) = 4.856; *p* = 0.030). No other variable was found to significantly influence log_2_ antibody titers over time (S2 Table).

### Booster vaccine I

Following Booster Vaccine I (BV1), 148 participants averaged a mean peak titer of 17.74 ± 1.14. Table 5 details significant differences observed across all extrinsic and intrinsic health factors in the BV1 sub-cohort or that were noted to be significant in the PV and BV2 sub-cohorts. Other measures are documented in S3 Table.

**Table 5.** Mean log_2_ Ab titers in first booster vaccination sub-cohort with varying extrinsic and intrinsic health factors.

EXTRINSIC HEALTH FACTORS – BOOSTER VACCINATION I
| ***COVID-19*** | N | Mean log<sub>2</sub> Ab (± SD) | *p* |
| --- | --- | --- | --- |
| *Booster Vaccination I (n = 98)* |  |  |  |
| *No* | 68 (69.39%) | 14.70 (± 1.20) | 0.649 |
| *Yes* | 30 (30.61%) | 14.81 (± 1.02) |  |
| ***Drug Use*** |  |  |  |
| *Booster Vaccination I (n=97)* |  |  |  |
| *No* | 93 (95.88%) | 14.74 (± 1.15) | 0.794 |
| *Yes* | 4 (4.12%) | 14.89 (± 0.96) |  |
| *INTRINSIC HEALTH FACTORS – VACCINATED PARTICIPANTS* |  |  |  |
| ***Age*** |  |  |  |
| *Booster Vaccination I (n=98)* |  |  |  |
| *<40 – Young* | 24 (24.49%) | 15.06 (± 0.97) | 0.277 |
| *40-65 – Middle* | 43 (43.88%) | 14.62 (± 0.96) |  |
| *>65 – Elderly* | 31 (31.63%) | 14.64 (± 1.44) |  |
| ***Biological Sex*** |  |  |  |
| *Booster Vaccination I (n=98)* |  |  |  |
| *Female* | **50 (51.02%)** | **14.96 (± 1.22)** | **0.043 \*** |
| *Male* | **48 (48.98%)** | **14.50 (± 1.01)** |  |
| ***Employment Status*** |  |  |  |
| *Booster Vaccination I (n=97)* |  |  |  |
| *Employed* | 78 (80.41%) | 14.82 (± 0.94) | 0.541 |
| *Retired* | 14 (14.43%) | 14.29 (± 1.91) |  |
| *Unemployed* | 5 (5.16%) | 15.04 (± 0.89) |  |
| ***Hispanic/Non-Hispanic*** |  |  |  |
| *Booster Vaccination I (n=98)* |  |  |  |
| *Hispanic* | 34 (34.69%) | 14.97 (± 0.98) | 0.144 |
| *Non-Hispanic* | 64 (65.31%) | 14.61 (± 1.21) |  |

Significant differences in peak Ab were observed between males ( $14.50 \pm 1.01$ ) and females
( $14.96 \pm 1.22$ ) following BV1 ( $T = -2.05$ ; $p = 0.043$ ).

#### Extrinsic health factors

Of the 95 participants that received BV1, 3 (3.16%) had CVD and 92 (96.84%) did not have CVD. Additionally, the mean log_2_ Ab of participants with CVD (14.64 ± 1.00) and participants without CVD (14.75 ± 1.15) did not significantly vary (*T* = 0.16; *p* = 0.872) after BV1. Three (3.12%) had diabetes and 93 (96.88%) did not, but diabetic (13.98 ± 1.53) and non-diabetic (14.77 ± 1.14) participants did not have observable differences (*T* = 1.19; *p* = 0.239) in peak titers. Of the 89 participants that received BV1, 23 (25.84%) had hypercholesterolemia and 66 (74.16%) did not. Eighteen (18.95%) participants had hypertension and 77 (81.05%) did not. Among participants with hypercholesterolemia (14.73 ± 0.95) or without (14.81 ± 1.25), and hypertension (14.81 ± 0.22) or without (14.75 ± 1.12), we did not observe differences in peak BV1 response (*T* = 0.28; *p* = 0.781, *T* = -0.21; *p* = 0.836).

Thirty (30.61%) participants who received BV1 were previously infected with SARS-CoV-2 and 68 (69.39%) were not. Furthermore, the mean log_2_ Ab of participants with a COVID-19 history (14.81 ± 1.02) and no history (14.70 ± 1.20) were not statistically different (*T* = -0.46; *p* = 0.649) after BV1. In the BV1 sub-cohort, 17 (17.52%) received a full-dose Moderna mRNA-1273 vaccine, while 24 (24.74%) received a half-dose Moderna mRNA-1273 vaccine, and 56 (56.74%) received a Pfizer BNT162b2 vaccine, but the mean log_2_ Ab in participants with full-dose Moderna mRNA-1273 (14.82 ± 1.55 ; *Tukey* = A), half-dose Moderna mRNA-1273 (15.14 ± 0.78 ; *Tukey* = A), and Pfizer BNT162b2 (14.54 ± 1.10 log_2_ Ab ; *Tukey* = A) did not have significantly distinct peak responses (*F* = 2.51 ; *p* = 0.087).

Interestingly, the Pfizer BNT162b2 BV1 group exhibited a very homogeneous peak response following vaccination (14.54; SE = 0.02) while ultimately it was not statistically different from the other groups. Of the 97 participants that received BV1, 74 (76.29%) were alcohol consumers and 23 (23.71%) were not. The peak response of alcohol consumers (14.85 ± 1.01) and non-consumers (14.43 ± 1.48) were not significantly different (*T* = -1.55; *p* = 0.124). Four (4.12%) participants were recreational drug users and 93 (95.88%) were not, and the peak response was not significantly different (*T* = -0.26; *p* = 0.794) between drug users (14.89 ± 0.96) and non-users (14.74 ± 1.15).

#### Intrinsic health factors

Twenty-four (24.49%) young participants were in the BV1 sub-cohort, while 43 (43.88%) were of middle age, and 34 (34.69%) were elderly. The peak response among different age groups was not significantly different (*F* = 1.30; *p* = 0.277), including young (15.06 ± 0.97; *Tukey* = A), middle-aged (14.62 ± 0.96, *Tukey* = A), and elderly participants (14.46 ± 1.44; *Tukey* = A). Of the 98 participants that received BV1, 50 (51.02%) were female and 48 (49.98%) were male, and a significantly lower peak response (*T* = -2.05; *p* = 0.043) was observed in males (14.50 ± 1.01) in comparison to females (14.96 ± 1.22).

Of the 98 participants that received BV1, 10 (10.20%) obtained an associate degree or technical degree, 21 (21.43%) a bachelor’s degree, 2 (2.04%) a high school diploma, 48 (48.98%) a master’s degree or higher, and 17 (17.35%) reported an unlisted level. The mean log_2_ Ab between participants with an associate degree or technical degree (14.84 ± 1.62; *Tukey* = A), a bachelor’s degree (14.64 ± 1.38; *Tukey* = A), a master’s degree or higher (14.81 ± 0.91; *Tukey* = A), and other educational levels (14.64 ± 1.23; *Tukey* = A) were not statistically different (*F* = 0.17; *p* = 0.916). Seventy-eight participants (80.41%) were employed, 14 (14.43%) were retired, and 5 (5.16%) were unemployed. We observed that the variances of log_2_ Ab between employed, retired and unemployed participants were statistically different (*F* (*2,147*) = 4.41; *p* = 0.015) after BV1, though the peak responses were not significantly different (*F* = 0.66; *p* = 0.541) between these groups (14.82 ± 0.94, *Games-Howell* = A; 14.29 ± 1.91, *Games-Howell* = A; 15.04 ± 0.89; *Games-Howell* = A; respectively).

In the BV1 sub-cohort, 8 (8.16%) participants were divorced, 4 (4.08%) participated in a domestic partnership, 61 (62.24%) were married, 22 (22.45%) were single, and 3 (3.07%) were widowed. The mean log_2_ Ab between divorced (15.02 ± 1.06; *Tukey* = A), domestic partnership (14.39 ± 0.96; *Tukey* = A), married (14.68 ± 1.22 ; *Tukey* = A), single (14.83 ± 1.01; *Tukey* = A), and widowed (14.98 ± 1.16; *Tukey* = A) participants were not statistically different (*F* = 0.31; *p* = 0.869) after BV1. One (1.02%) identified as bisexual, 6 (6.12%) identified as gay, 90 (91.84%) identified as heterosexual, and 1 identified as lesbian (1.02%), but the mean log_2_ Ab between gay (14.31 ± 0.82) and heterosexual (14.74 ± 1.15) participants were not significantly different (*T* = -0.90; *p*=0.368) following BV1.

Finally, of the 98 participants that received BV1 8 (8.16%) were Asian, 3 (3.06%) were African American, 5 (5.11%) identified under another race, and 82 (83.67%) were White. The peak responses between participants among different races were not statistically different (*F* = 0.41; *p* = 0.747), including Asian (14.77 ± 0.84 ; *Tukey* = A), African American (15.32 ± 0.58; *Tukey* = A), Other (15.04 ± 0.55 ; *Tukey* = A), and White (14.69 ± 1.21; *Tukey* = A). Of the 98 participants that received BV1, 34 (34.69%) identified as Hispanic, and 64 (65.31%) identified as non-Hispanic. The mean log_2_ Ab between Hispanic (14.97 ± 0.98) and non-Hispanic (14.61 ± 1.21) participants were not statistically different (*T* = 1.47; *p* = 0.144) after BV1.

#### Longitudinal analysis following booster vaccination I

Linear mixed effects models were generated to examine the effect of IHFs and EHFs on log_2_ antibody titers over time following BV1 (Fig 6). Overall, we found that the quadratic (*estimate* = -6.657e-04; *F*(1, 72) = 4.007; *p* = 0.049) and cubic (*estimate* = 1.827e-06; *F*(1, 72) = 10.142; *p* = 0.002) terms for days elapsed since the first booster were observed to be a significant predictor of log_2_ antibody titers over time, elucidating a non-linear decay pattern following BV1. Curiously, this analysis also revealed that drug use positively impacted log_2_ antibody titers, (*estimate* = 2.527e+00; *F*(1, 24) = 8.917; *p =* .006). Following post-hoc testing, booster type (i.e., Pfizer BNT162b2 vs. Moderna mRNA-1273 (full dose) vs. Moderna mRNA-1273 (half dose) was found to significantly affect the trajectory of log_2_ antibody titers, (*estimate* = 3.997416e-01; *F*(2, 24) = 4.*755*; *p* = .018), with those who received a half-dose of the Moderna mRNA-1273 booster displaying increased log_2_ antibody titers boasting significantly higher titers than who received the Pfizer BNT162b2 booster.

**Fig 6.**
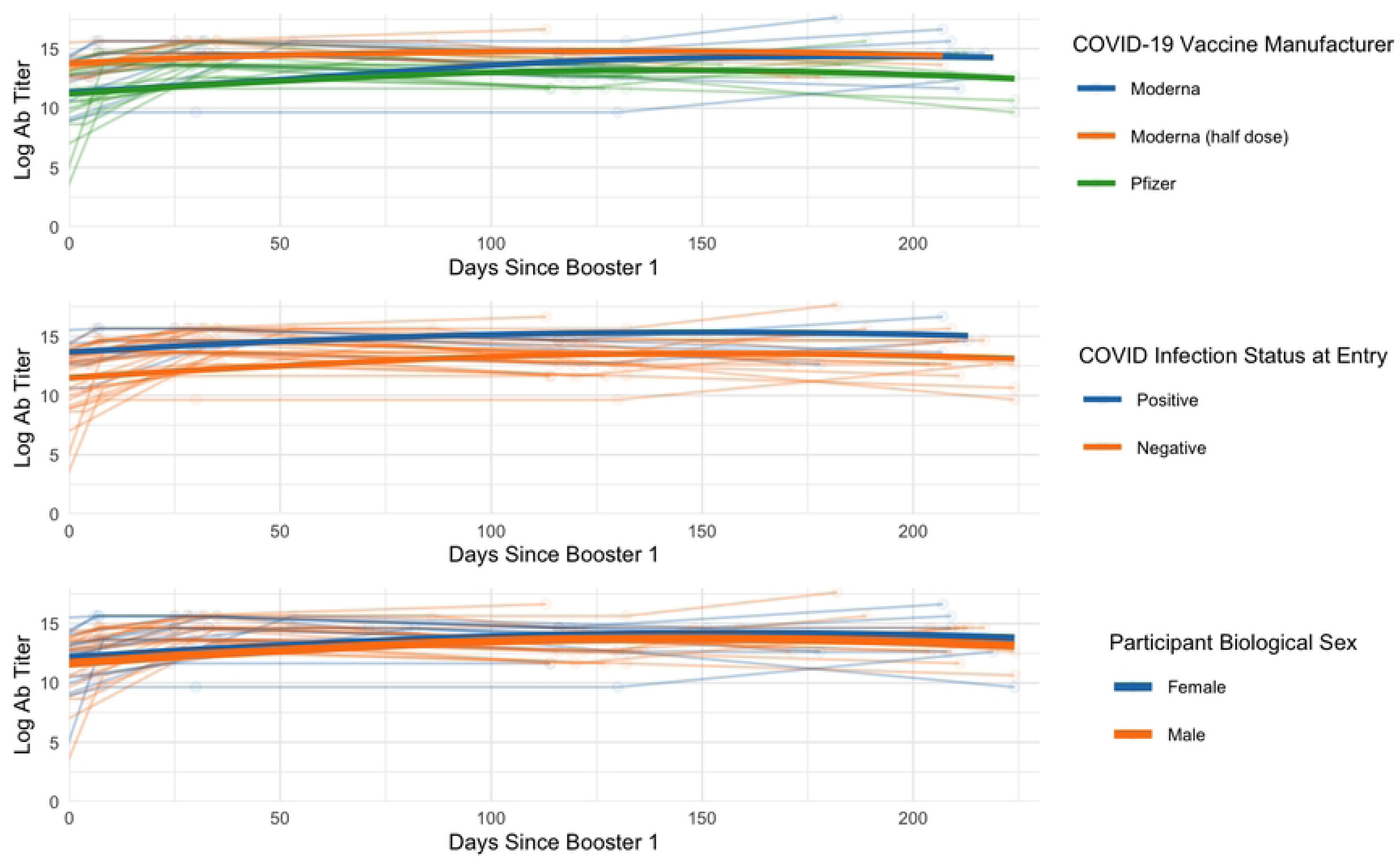
Linear mixed-effects model for investigation of IHFs and EHFs on log_2_ antibody titers over time following BV1. The quadratic and cubic terms for days indicate a nonlinear decay of titers over time. Additionally, the model indicated that drug use and vaccination with the half-dose Moderna mRNA-1273 vaccine positively impacted titers. Other factors did not contribute significantly to titers over time.

Notably, factors such as ethnicity, gender, age at entry, race, and the linear term for the number of days since the first boost did not significantly contribute to the variation in log_2_ antibody titers, all *p* > .05 (S4 Table).

### Booster vaccine II

In the cohort, 24 participants received BV2 and averaged 15.06 ± 1.61 Ab following vaccination. Table 6 details the extrinsic and intrinsic health factors that observed a significant difference within groups in the BV2 cohort or that had significance in the PV and BV1 sub-cohorts. Other measures are documented in S5 Table.

**Table 6.**
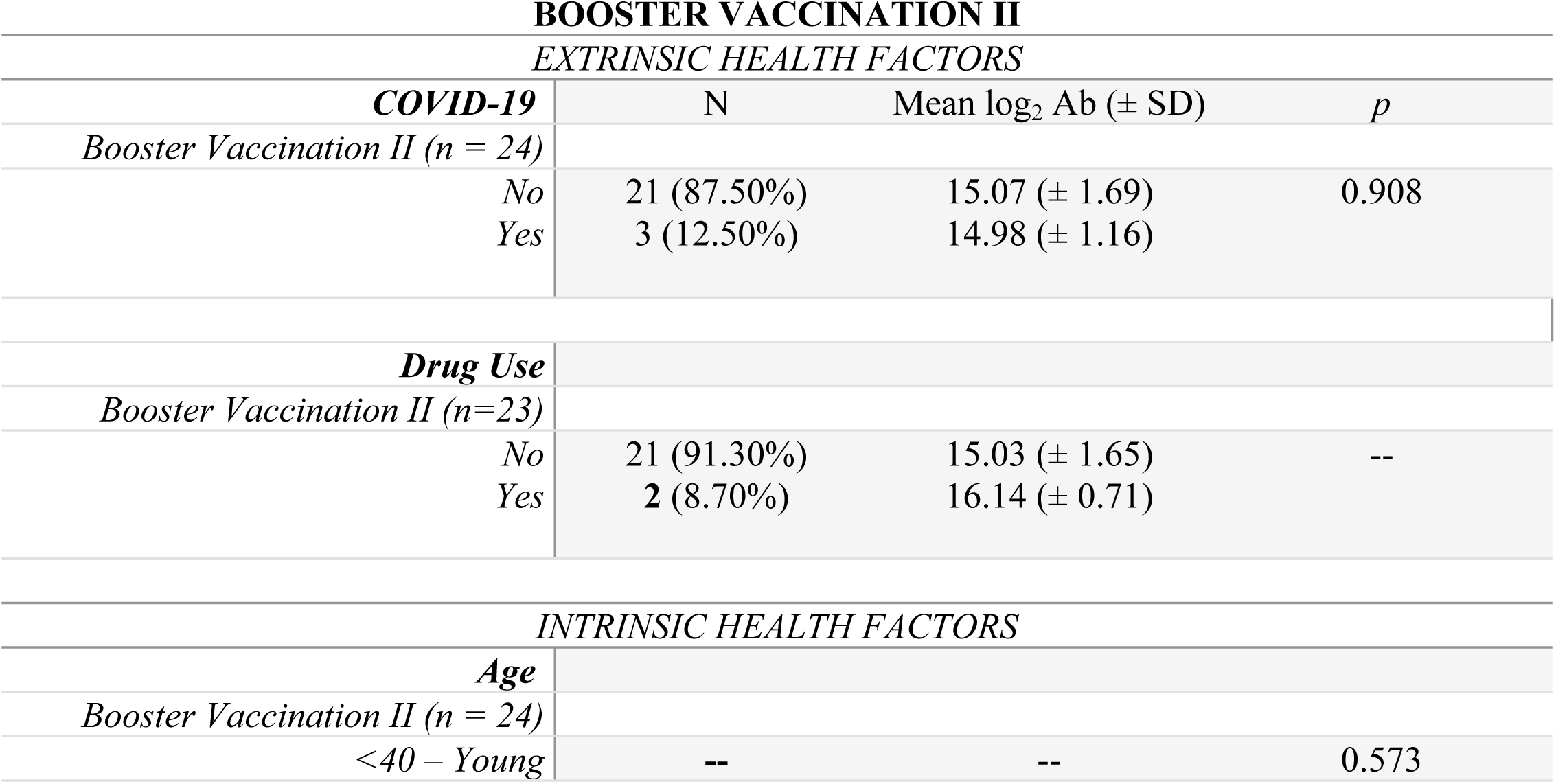
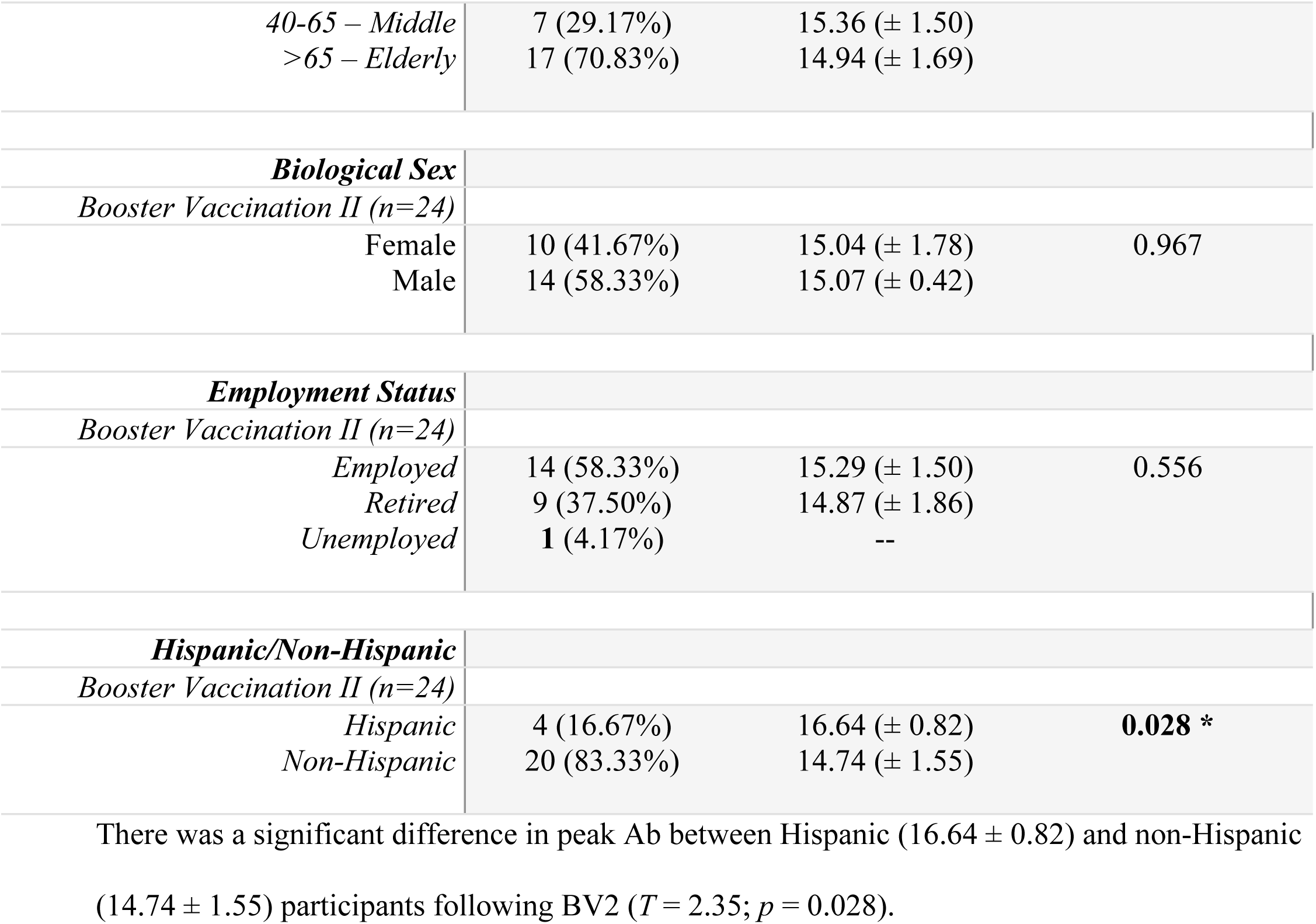
Mean log_2_ Ab titers in second booster vaccination sub-cohort with varying extrinsic and intrinsic health factors.

#### Extrinsic health factors

In the BV2 sub-cohort 2 (9.09%) had CVD and 20 (90.91%) did not have CVD, 2 (8.70%) had diabetes and 21 (91.30%) did not have diabetes, and 10 (47.62%) had hypercholesterolemia and 11 (52.38%) did not have hypercholesterolemia. The peak response of participants with hypercholesterolemia (15.54 ± 0.99) and without (14.92 ± 2.01) were not statistically different (*T* = -0.89; *p* = 0.383) after BV2. Five (21.74%) had hypertension and 18 (78.26%) did not, but the peak responses between those with hypertension (15.84 ± 0.37) and without (14.92 ± 1.74) were not statistically different (*T* = -1.13; *p* = 0.270).

Three (12.50%) participants from the BV2 sub-cohort were infected with SARS-CoV-2 prior to enrollment and 21 (87.50%) were not. The mean log_2_ Ab of participants with prior SARS-CoV-2 infection (14.98 ± 1.16) and no prior infection (15.07 ± 1.69) were not statistically significant (*T* = 0.13; *p* = 0.908). Thirteen (54.17%) received a full-dose Moderna mRNA-1273 vaccine, while 11 (45.83%) received a Pfizer BNT162b2 vaccine, but the peak responses between Moderna mRNA-1273 (15.41 ± 1.74) and Pfizer BNT162b2-vaccinated (14.64 ± 1.41) participants were not significantly different (*T* = 1.17; *p* = 0.253). Of the 23 participants that received BV2, 17 (73.91%) were alcohol consumers and 6 (26.09%) were not. The mean log_2_ Ab of alcohol consumers (14.94 ± 1.21) and non-alcohol consumers (15.64 ± 2.53) were not significantly different (*T* = 0.91; *p* = 0.371) after BV2. Two (8.70%) participants were drug users and 21 (91.30%) were not.

#### Intrinsic health factors

Of the 24 participants that received BV2, 7 (29.17%) were middle aged and 17 (70.83%) were elderly. The peak response of middle-aged participants (15.36 ± 1.50) and elderly participants (14.96 ± 1.69) was not significantly different (*T* = -0.57; *p* = 0.573). Ten (41.67%) participants were female and 14 (58.33%) were male, and the peak response in male (15.07 ± 0.42) and female (15.04 ± 1.78) participants was not statistically different (*T* = -0.04; *p* = 0.967) after BV2.

Two (8.33%) participants had an associate degree or technical degree, 5 (20.83%) had a bachelor’s degree, 13 (54.17%) had a master’s degree or higher, and 4 (16.67%) had an unlisted degree. The peak responses among participants in this group were not statistically different (*F* = 0.64, *p* = 0.539), including those with a bachelor’s degree (14.44 ± 2.39; *Tukey* = A), a master’s degree or higher (14.03 ± 1.26; *Tukey* = A), and other educational levels (15.64 ± 1.41; *Tukey*=A). Of the 24 participants that received BV2, 14 (58.33%) were employed, 9 (37.50%) were retired, and 1 (4.17%) was unemployed. The Ab of employed (15.29 ± 1.50) and retired (14.87 ± 1.86) participants were not statistically different (*T* = 0.60; *p* = 0.556) after BV2.

In the BV2 cohort, 3 (12.50%) participants were divorced, 1 (4.17%) participated in a domestic partnership, 16 (66.67%) were married, 2 (8.33%) were single, and 2 (8.33%) were widowed. The mean log_2_ Ab between divorced (15.98 ± 0.58) and married (14.77 ± 1.75) participants were not statistically different (*T* = 1.61; *p* = 0.261). Two (8.33%) identified as gay and 22 (91.67%) identified as heterosexual.

Four (16.67%) identified as Hispanic, and 20 (83.33%) identified as non-Hispanic. The peak response between Hispanic (16.64 ± 0.82) and non-Hispanic (14.74 ± 1.55) participants were statistically different (*T* = 2.35; *p* = 0.028) after BV2. Of the 24 participants that received BV2, 2 (8.33%) were Asian, 2 (8.33%) were African American, 2 (8.33%) identified under another race, and 18 (75.01%) were White.

#### Longitudinal analysis following booster vaccination II

Lastly, linear mixed effects models were used to generate a growth curve of the log_2_ Ab response following BV2. Again, the quadratic term for the number of days since the 2^nd^ booster emerged as a significant predictor of log_2_ antibody titers, (*estimate* = -6.628^e-05; *F*(1, 104) = 7.916; *p* = .006), suggestive of a non-linear decay pattern following vaccination. No other covariate of interest was found to significantly affect the trajectory of Ab over time, including linear or cubic time, gender, booster type, or drug use (S6 Table). Overall, these results suggest that a 4^th^ dose of an mRNA COVID-19 vaccine provide an additive effect on the immune response, dampening the role of underlying IHF and EHFs that may influence the post-vaccine immune response.

## Discussion

In this work, we sought to evaluate the effects of intrinsic and extrinsic health factors on the peak antibody response as well as characterize antibody trajectory over time following COVID-19 mRNA primary vaccination and booster vaccination. Through cross-sectional analysis, we found that EHFs influenced Ab production after primary vaccination. Participants that reported recreational drug use, including marijuana, cocaine, and amphetamines had marginally higher log_2_ Ab compared to non-users after PV, but not BV1 or BV2. Previous work has shown that recreational drug use is immunomodulatory, increasing the susceptibility to infectious diseases and microbes while impairing antibody production following vaccination (30–33). Our findings support the opposite, though this warrants further investigation given the small sample size of drug users in our cohort. This was further substantiated in linear mixed-effects models, which demonstrated that participants with reported drug use had higher titers over time only after BV1.

Our cross-sectional analyses were inherently limited in their ability to control for all other variables that may influence humoral responses and were utilized in the context of this work to capture only the peak antibody response. Further, unlike mixed effects models, a cross-sectional approach does not permit the assessment of the impact of individual covariates while controlling for the effect of time elapsed since vaccination. We also found that individuals without hypertension had a lower and more homogeneous Ab response following PV, but not BV1 or BV2, which may be attributed to previously reported changes in T cell immunometabolism (i.e., aberrant T cell activation, differentiation, and proliferation) (34–38). Studies evaluating the effect of hypertension on antibody titers have provided variable results, with some supporting hypertension as a factor driving vaccine immunogenicity (37, 38), and others refuting it (39). We also observed significantly higher mean peak log_2_ Ab in participants with previous SARS-CoV-2 infection compared to those without a history after only PV, consistent with previous findings (5, 40, 41). Lastly, no significant difference in the peak log_2_ Ab was found between participants that received Moderna mRNA-1273 and Pfizer BNT162b2 vaccines after vaccination, although this was not the case over time.

Young (low age) individuals had higher peak Ab compared to elderly participants, but not middle-aged participants only after PV, which may be attributed to age-related defects in lymphocytes like mature lymphocyte senescence or decreased production and proliferation of lymphocytes and their associated biomarkers (42–46). Furthermore, we observed significantly lower Ab in retired participants following PV, which likely corresponds to the advanced age of those participants. Also, we found that non-Hispanic participants had lower Ab than Hispanic after BV2, but our sample size is limited and therefore may be limited in generalizability. Lastly, we found that females had significantly higher Ab following PV and BV1, but not BV2. Differences in immune responses based on biological sex may be linked to phylogenetic preservation and subsequent reproductive success associated with more robust humoral immune reaction in females (47). Accordingly, the decreased peak Ab responses we observed in males is supported by previously documented sex-related humoral and cellular-mediated differences (40, 47–49).

The overall findings from the cross-sectional approach indicate, at least in part, the role of EHFs and IHFs in generating differential humoral immunes response following COVID-19 vaccination. Notably, these analyses did not control for other variables, which increases the likelihood of introducing confounding factors in the analysis. Thus, cross-sectional analyses should be considered useful in identifying differences in immune response following vaccination but are perhaps limited in evaluating how these change over time. Additionally, controlling for other factors can help determine more robust relationships between Ab and IHFs/EHFs. Because the immune system follows a dynamic response following activation both through time and due to inter-individual differences, adopting models that account for this and can control for these differences may be used to corroborate cross-sectional analysis. Thus, distinct linear mixed-effects models were used to investigate the factors influencing the log-transformed titer levels following primary and booster vaccination. The number of days elapsed since primary vaccination was negatively associated with antibody titer magnitude, suggesting that as time passes since full vaccination, the titer levels decrease. This time-dependent decay of antibodies was observed to have significant nonlinear components, particularly following BV1 and BV2.

We also found that COVID-19 positive individuals had higher antibody levels following primary vaccination compared to COVID-19 negative individuals, and male participants had lower antibody titers compared to females. The type of vaccine received also affected antibody magnitude and durability; those who received the Pfizer BNT162b2 vaccine had lower titers than those who were given the Moderna mRNA-1273 vaccine. Differential antibody production between Moderna mRNA-1273 and Pfizer BNT162b2 vaccines observed in our cohort are consistent with previous findings, which could be due to varied anti-spike activity generated by each vaccine type (50, 51). We also observed that Moderna mRNA-1273 half-dose recipients had higher post-booster Ab compared to those who received Pfizer BNT162b2 vaccination over time, but this effect was not observed following BV2. Interestingly, post-BV2 Ab were not significantly affected by any predictive factors. This lack of predictors, including vaccine manufacturer, suggest a more robust and homogeneous humoral immune response following BV2.

## Conclusions

Overall, this study aimed to examine the effects of intrinsic and extrinsic health factors on Ab following vaccination and through time using cross-sectional and longitudinal models. Differences in antibody titers among the cohort were observed after primary vaccination for individuals with a history of drug use, prior SARS-CoV-2 infection, hypertension, age, and biological sex. Differential titer production was also observed across females following BV1 and Hispanic participants following BV2. Linear-mixed effects models used in this study aimed to control for random and fixed effects like population responses and individual Ab variations over time and revealed trends in the decline of Ab over time following full vaccination, as well as effects by COVID-19 vaccine manufacturer, prior SARS-CoV-2 infection, and biological sex. Drug users and half-dose Moderna mRNA-1273 recipients had higher peak antibody titers over time following the first booster, while no predictive factors significantly affected post-second booster antibody responses. The absence of predictive factors for second booster immunogenicity suggests a more robust and consistent immune response after the second booster vaccine administration.

## Data Availability

All relevant de-identified data is available upon request.

## Acknowledgments

We thank Margaret Roach, Elizabeth Varghese, Maria Pallin, Celeste Sanchez, and Ailet Reyes for their technical contributions. We also thank Aria Nawab and Daniel Muniz for their assistance in sample collection, and Alexander Kizhner for his assistance with data management and in generating the early statistical analysis.

## Supporting information

**S1 Table. Other Measured Extrinsic and Intrinsic Health Factors Among the Primary Vaccination Sub-Cohort. ǂǂǂǂ: Levene’s test found a statistically significant difference in the variances between participants with and without hypertension (F** (**1,145**) **=8.15, p=0.005).**

**S2 Table. Linear Mixed Effects Model (LMM) Evaluating the Relationship Between Post-Primary Vaccination Antibody Titers and Time, COVID-19 Vaccine Manufacturer, Prior COVID-19 Infection Status, and Biological Sex.**

**S3 Table. Other Measured Extrinsic and Intrinsic Health Factors Among the First Booster Vaccination Sub-Cohort.**

**S4 Table. Linear Mixed Effects Model (LMM) Evaluating the Relationship Between BV1 Antibody Titers and Time, COVID-19 Vaccine Manufacturer, Prior COVID-19 Infection Status, and Biological Sex.**

**S5 Table. Other Measured Extrinsic and Intrinsic Health Factors Among the Second Booster Vaccination Sub-Cohort.**

**S6 Table. Linear Mixed Effects Model (LMM) Evaluating the Relationship Between BV2 Antibody Titers and Time, COVID-19 Vaccine Manufacturer, Prior COVID-19 Infection Status, and Biological Sex.**

## References

1. Wendisch D, Dietrich O, Mari T, von Stillfried S, Ibarra IL, Mittermaier M, et al. SARS-CoV-2 infection triggers profibrotic macrophage responses and lung fibrosis. Cell. 2021;184(26):6243–61.e27.

2. Jacobs JL, Haidar G, Mellors JW. COVID-19: Challenges of Viral Variants. Annu Rev Med. 2023;74:31–53.

3. Shishido AA, Barnes AH, Narayanan S, Chua JV. COVID-19 Vaccines-All You Want to Know. Semin Respir Crit Care Med. 2023;44(1):143–72.

4. Tsai SC, Lu CC, Bau DT, Chiu YJ, Yen YT, Hsu YM, et al. Approaches towards fighting the COVID-19 pandemic (Review). Int J Mol Med. 2021;47(1):3–22.

5. Williams EC, Kizhner A, Stark VS, Nawab A, Muniz DD, Echeverri Tribin F, et al. Predictors for reactogenicity and humoral immunity to SARS-CoV-2 following infection and mRNA vaccination: A regularized, mixed-effects modelling approach. Frontiers in Immunology. 2023;14.

6. Moshkovits I, Shepshelovich D. Emergency Use Authorizations of COVID-19-Related Medical Products. JAMA Intern Med. 2022;182(2):228–9.

7. Parums DV. Editorial: First Full Regulatory Approval of a COVID-19 Vaccine, the BNT162b2 Pfizer-BioNTech Vaccine, and the Real-World Implications for Public Health Policy. Med Sci Monit. 2021;27:e934625.

8. Forchette L, Sebastian W, Liu T. A Comprehensive Review of COVID-19 Virology, Vaccines, Variants, and Therapeutics. Curr Med Sci. 2021;41(6):1037–51.

9. Callegaro A, Borleri D, Farina C, Napolitano G, Valenti D, Rizzi M, et al. Antibody response to SARS-CoV-2 vaccination is extremely vivacious in subjects with previous SARS-CoV-2 infection. J Med Virol. 2021;93(7):4612–5.

10. Crawford KHD, Dingens AS, Eguia R, Wolf CR, Wilcox N, Logue JK, et al. Dynamics of Neutralizing Antibody Titers in the Months After Severe Acute Respiratory Syndrome Coronavirus 2 Infection. J Infect Dis. 2021;223(2):197–205.

11. Hall V, Foulkes S, Insalata F, Kirwan P, Saei A, Atti A, et al. Protection against SARS-CoV-2 after Covid-19 Vaccination and Previous Infection. N Engl J Med. 2022;386(13):1207–20.

12. Kelsen SG, Braverman AS, Aksoy MO, Hayman JA, Patel PS, Rajput C, et al. SARS-CoV-2 BNT162b2 vaccine-induced humoral response and reactogenicity in individuals with prior COVID-19 disease. JCI Insight. 2022;7(4).

13. Lozano-Ojalvo D, Camara C, Lopez-Granados E, Nozal P, Del Pino-Molina L, Bravo-Gallego LY, et al. Differential effects of the second SARS-CoV-2 mRNA vaccine dose on T cell immunity in naive and COVID-19 recovered individuals. Cell Rep. 2021;36(8):109570.

14. Mungmunpuntipantip R, Wiwanitkit V. Antibody response to SARS-CoV-2 vaccination, previous SARS-CoV-2 infection, and change to single-dose vaccination. J Med Virol. 2021;93(12):6474.

15. Nagura-Ikeda M, Imai K, Kubota K, Noguchi S, Kitagawa Y, Matsuoka M, et al. Clinical characteristics and antibody response to SARS-CoV-2 spike 1 protein using VITROS Anti-SARS-CoV-2 antibody tests in COVID-19 patients in Japan. J Med Microbiol. 2021;70(4).

16. Ontañón J, Blas J, de Cabo C, Santos C, Ruiz-Escribano E, García A, et al. Influence of past infection with SARS-CoV-2 on the response to the BNT162b2 mRNA vaccine in health care workers: Kinetics and durability of the humoral immune response. EBioMedicine. 2021;73:103656.

17. Pereson MJ, Amaya L, Neukam K, Baré P, Echegoyen N, Badano MN, et al. Heterologous gam-COVID-vac (sputnik V)/mRNA-1273 (moderna) vaccination induces a stronger humoral response than homologous sputnik V in a real-world data analysis. Clinical Microbiology and Infection. 2022;28(10):1382–8.

18. Tsang JS, Dobaño C, VanDamme P, Moncunill G, Marchant A, Othman RB, et al. Improving Vaccine-Induced Immunity: Can Baseline Predict Outcome? Trends Immunol. 2020;41(6):457–65.

19. Young KM, Gray CM, Bekker L-G. Is Obesity a Risk Factor for Vaccine Non-Responsiveness? PLOS ONE. 2013;8(12):e82779.

20. Boroumand AB, Forouhi M, Karimi F, Moghadam AS, Naeini LG, Kokabian P, et al. Immunogenicity of COVID-19 vaccines in patients with diabetes mellitus: A systematic review. Frontiers in Immunology. 2022;13.

21. Flanagan KL, van Crevel R, Curtis N, Shann F, Levy O, Network ftO. Heterologous (“Nonspecific”) and Sex-Differential Effects of Vaccines: Epidemiology, Clinical Trials, and Emerging Immunologic Mechanisms. Clinical Infectious Diseases. 2013;57(2):283–9.

22. Posteraro B, Pastorino R, Di Giannantonio P, Ianuale C, Amore R, Ricciardi W, et al. The link between genetic variation and variability in vaccine responses: Systematic review and meta-analyses. Vaccine. 2014;32(15):1661–9.

23. Grubeck-Loebenstein B, Della Bella S, Iorio AM, Michel J-P, Pawelec G, Solana R. Immunosenescence and vaccine failure in the elderly. Aging Clinical and Experimental Research. 2009;21(3):201–9.

24. Kumar R, Burns EA. Age-related decline in immunity: implications for vaccine responsiveness. Expert Review of Vaccines. 2008;7(4):467–79.

25. Simon V, Kota V, Bloomquist RF, Hanley HB, Forgacs D, Pahwa S, et al. PARIS and SPARTA: Finding the Achilles’ Heel of SARS-CoV-2. mSphere. 2022;7(3):e0017922.

26. HANC. Cross-Network PBMC Processing Standard Operating Procedure. 2018. p. 1–45.

27. Stadlbauer D, Amanat F, Chromikova V, Jiang K, Strohmeier S, Arunkumar GA, et al. SARS-CoV-2 Seroconversion in Humans: A Detailed Protocol for a Serological Assay, Antigen Production, and Test Setup. Curr Protoc Microbiol. 2020;57(1):e100.

28. Amanat F, Stadlbauer D, Strohmeier S, Nguyen THO, Chromikova V, McMahon M, et al. A serological assay to detect SARS-CoV-2 seroconversion in humans. Nat Med. 2020;26(7):1033–6.

29. Sette A, Crotty S. Immunological memory to SARS-CoV-2 infection and COVID-19 vaccines. Immunol Rev. 2022;310(1):27–46.

30. Baral S, Sherman SG, Millson P, Beyrer C. Vaccine immunogenicity in injecting drug users: a systematic review. Lancet Infect Dis. 2007;7(10):667–74.

31. Friedman H, Newton C, Klein TW. Microbial infections, immunomodulation, and drugs of abuse. Clin Microbiol Rev. 2003;16(2):209–19.

32. Friedman H, Pross S, Klein TW. Addictive drugs and their relationship with infectious deseases. FEMS Immunology & Medical Microbiology. 2006;47(3):330–42.

33. Re G-F, Jia J, Xu Y, Zhang Z, Xie Z-R, Kong D, et al. Dynamics and correlations in multiplex immune profiling reveal persistent immune inflammation in male drug users after withdrawal. International Immunopharmacology. 2022;107:108696.

34. Mattson DL, Abais-Battad JM. T Cell Immunometabolism and Redox Signaling in Hypertension. Curr Hypertens Rep. 2021;23(12):45.

35. Moshfegh CM, Case AJ. The Redox-Metabolic Couple of T Lymphocytes: Potential Consequences for Hypertension. Antioxid Redox Signal. 2021;34(12):915–35.

36. Rai A, Narisawa M, Li P, Piao L, Li Y, Yang G, et al. Adaptive immune disorders in hypertension and heart failure: focusing on T-cell subset activation and clinical implications. J Hypertens. 2020;38(10):1878–89.

37. Soegiarto G, Wulandari L, Purnomosari D, Dhia Fahmita K, Ikhwan Gautama H, Tri Hadmoko S, et al. Hypertension is associated with antibody response and breakthrough infection in health care workers following vaccination with inactivated SARS-CoV-2. Vaccine. 2022;40(30):4046–56.

38. Watanabe M, Balena A, Tuccinardi D, Tozzi R, Risi R, Masi D, et al. Central obesity, smoking habit, and hypertension are associated with lower antibody titres in response to COVID-19 mRNA vaccine. Diabetes Metab Res Rev. 2022;38(1):e3465.

39. Pellini R, Venuti A, Pimpinelli F, Abril E, Blandino G, Campo F, et al. Initial observations on age, gender, BMI and hypertension in antibody responses to SARS-CoV-2 BNT162b2 vaccine. EClinicalMedicine. 2021;36:100928.

40. Wolszczak-Biedrzycka B, Bieńkowska A, Dorf J. Assessment of Post-Vaccination Antibody Response Eight Months after the Administration of BNT1622b2 Vaccine to Healthcare Workers with Particular Emphasis on the Impact of Previous COVID-19 Infection. Vaccines. 2021;9(12):1508.

41. Krammer F, Srivastava K, Alshammary H, Amoako AA, Awawda MH, Beach KF, et al. Antibody Responses in Seropositive Persons after a Single Dose of SARS-CoV-2 mRNA Vaccine. New England Journal of Medicine. 2021;384(14):1372–4.

42. Bajaj V, Gadi N, Spihlman AP, Wu SC, Choi CH, Moulton VR. Aging, Immunity, and COVID-19: How Age Influences the Host Immune Response to Coronavirus Infections? Frontiers in Physiology. 2021;11.

43. Gustafson CE, Kim C, Weyand CM, Goronzy JJ. Influence of immune aging on vaccine responses. Journal of Allergy and Clinical Immunology. 2020;145(5):1309–21.

44. Song H, Price PW, Cerny J. Age-related changes in antibody repertoire: contribution from T cells. Immunological Reviews. 1997;160(1):55–62.

45. Weksler ME. Changes in the B-cell repertoire with age. Vaccine. 2000;18(16):1624–8.

46. Montecino-Rodriguez E, Berent-Maoz B, Dorshkind K. Causes, consequences, and reversal of immune system aging. J Clin Invest. 2013;123(3):958–65.

47. Fink AL, Klein SL. The evolution of greater humoral immunity in females than males: implications for vaccine efficacy. Curr Opin Physiol. 2018;6:16–20.

48. Klein SL, Flanagan KL. Sex differences in immune responses. Nature Reviews Immunology. 2016;16(10):626–38.

49. Ortona E, Pierdominici M, Rider V. Editorial: Sex Hormones and Gender Differences in Immune Responses. Front Immunol. 2019;10:1076.

50. Kelliher MT, Levy JJ, Nerenz RD, Poore B, Johnston AA, Rogers AR, et al. Comparison of Symptoms and Antibody Response Following Administration of Moderna or Pfizer SARS-CoV-2 Vaccines. Arch Pathol Lab Med. 2022;146(6):677–85.

51. Keshavarz B, Richards NE, Workman LJ, Patel J, Muehling LM, Canderan G, et al. Trajectory of IgG to SARS-CoV-2 After Vaccination With BNT162b2 or mRNA-1273 in an Employee Cohort and Comparison With Natural Infection. Frontiers in Immunology. 2022;13.

